# Suppressive myeloid cells are a hallmark of severe COVID-19

**DOI:** 10.1101/2020.06.03.20119818

**Authors:** Jonas Schulte-Schrepping, Nico Reusch, Daniela Paclik, Kevin Baßler, Stephan Schlickeiser, Bowen Zhang, Benjamin Krämer, Tobias Krammer, Sophia Brumhard, Lorenzo Bonaguro, Elena De Domenico, Daniel Wendisch, Martin Grasshoff, Theodore S. Kapellos, Michael Beckstette, Tal Pecht, Adem Saglam, Oliver Dietrich, Henrik E. Mei, Axel R. Schulz, Claudia Conrad, Désirée Kunkel, Ehsan Vafadarnejad, Cheng-Jian Xu, Arik Horne, Miriam Herbert, Anna Drews, Charlotte Thibeault, Moritz Pfeiffer, Stefan Hippenstiel, Andreas Hocke, Holger Müller-Redetzky, Katrin-Moira Heim, Felix Machleidt, Alexander Uhrig, Laure Bousquillon de Jarcy, Linda Jürgens, Miriam Stegemann, Christoph R. Glösenkamp, Hans-Dieter Volk, Christine Goffinet, Jan Raabe, Kim Melanie Kaiser, Michael To Vinh, Gereon Rieke, Christian Meisel, Thomas Ulas, Matthias Becker, Robert Geffers, Martin Witzenrath, Christian Drosten, Norbert Suttorp, Christof von Kalle, Florian Kurth, Kristian Händler, Joachim L. Schultze, Anna C Aschenbrenner, Yang Li, Jacob Nattermann, Birgit Sawitzki, Antoine-Emmanuel Saliba, Leif Erik Sander, Deutsche COVID-19 OMICS Initiative (DeCOI)

## Abstract

‘Severe Acute Respiratory Syndrome - Coronavirus-2’ (SARS-CoV-2) infection causes Coronavirus Disease 2019 (COVID-19), a mild to moderate respiratory tract infection in the majority of patients. A subset of patients, however, progresses to severe disease and respiratory failure with acute respiratory distress syndrome (ARDS). Severe COVID-19 has been associated with increased neutrophil counts and dysregulated immune responses. The mechanisms of protective immunity in mild forms and the pathogenesis of dysregulated inflammation in severe courses of COVID-19 remain largely unclear. Here, we combined two single-cell RNA-sequencing technologies and single-cell proteomics in whole blood and peripheral blood mononuclear cells (PBMC) to determine changes in immune cell composition and activation in two independent dual-center patient cohorts (n=46+n=54 COVID-19 samples), each with mild and severe cases of COVID-19. We observed a specific increase of HLA-DR^hi^CD11c^hi^ inflammatory monocytes that displayed a strong interferon (IFN)-stimulated gene signature in patients with mild COVID-19, which was absent in severe disease. Instead, we found evidence of emergency myelopoiesis, marked by the occurrence of immunosuppressive pre-neutrophils and immature neutrophils and populations of dysfunctional and suppressive mature neutrophils, as well as suppressive HLA-DR^to^ monocytes in severe COVID-19. Our study provides detailed insights into systemic immune response to SARS-CoV-2 infection and it reveals profound alterations in the peripheral myeloid cell compartment associated with severe courses of COVID-19.

## Introduction

The emergence of severe acute respiratory syndrome coronavirus 2 (SARS-CoV-2) in December of 2019 (Wu et al., 2020) and the subsequent pandemic spread of Coronavirus Disease 2019 (COVID-19) has caused immense morbidity and mortality around the world (Fauver et al., 2020; Zhou et al., 2020b). Clinical presentations of COVID-19 are variable, and while the majority of patients experiences mild to moderate symptoms, a subset of 10-20% of patients develops pneumonia and severe disease (Brignola et al., 1988; Guan et al., 2020a; Huang et al., 2020; Wang et al., 2020b; Zhou et al., 2020a). Clinical deterioration and development of respiratory failure and acute respiratory distress syndrome (ARDS), typically develops in the second week of disease. Besides protracted viral replication, this kinetic suggests a role for secondary immune responses in the development of severe COVID-19 (Ziying Ong et al., 2020). However, the exact mechanisms that govern the pathophysiology of the different disease courses of COVID-19 remain ill-defined. Patients with comorbidities, including hypertension, diabetes, COPD, cardiovascular disease, and cerebrovascular disease are at highest risk to develop severe COVID-19 (Guan et al., 2020b; Wang et al., 2020a). Given that these conditions are associated with chronic inflammation, disease severity of COVID-19 might be closely linked to the underlying specific and nonspecific immune response to the virus.

SARS-CoV-2 was identified as the causative agent of COVID-19 (Wu et al., 2020) and similar to SARS coronavirus, it uses ACE2 as the primary cellular entry receptor (Hoffmann et al., 2020; Li et al., 2003). SARS-CoV-2 has a tropism for the upper airways and the lung (Wölfel et al., 2020), despite rather low numbers of cells that co-express ACE2 and the essential cofactor for ACE2 binding, TMPRSS2 (Allan et al., 2020; Qi et al., 2020; Sungnak et al., 2020; Ziegler et al., 2020), but the expression of ACE2 and TMPRSS2 in airway epithelial cells are increased by type-I IFN stimulation (Ziegler et al., 2020). Single cell studies of bronchoalveolar lavage samples have suggested a complex dysregulation of the pulmonary immune response in severe COVID-19 (Liao et al., 2020). Overall, systemic inflammation is linked to an unfavorable clinical course of disease and the development of severe COVID-19 (Giamarellos-Bourboulis et al., 2020; Ziying Ong et al., 2020). SARS-CoV2 infection induces specific T cell and B cell responses, which is reflected by elevation of SARS-CoV-2 peptide-specific T cells (Braun et al., 2020; Grifoni et al., 2020) and the production of SARS-CoV-2 specific antibody responses (Long et al., 2020; Ni et al., 2020). Patients with severe COVID-19 have high systemic levels of inflammatory cytokines, specifically IL-1β and IL-6 (Chen et al., 2020; Giamarellos-Bourboulis et al., 2020; Ziying Ong et al., 2020), whereas interferon (IFN) responses appear to be impaired as shown by whole blood transcriptomics (Hadjadj et al., 2020). Clinical observations and several studies indicate an increase of neutrophils and a decrease of non-classical (CD14^lo^CD16^hi^) monocytes in severe COVID-19 (Hadjadj et al., 2020; Merad and Martin, 2020). Severe immune dysregulation is a common phenomenon in sepsis, characterized by a progression from hyperinflammatory states to immunosuppression (Remy et al., 2020; Ritchie and Singanayagam, 2020) and similar mechanisms have been proposed for severe COVID-19 (Giamarellos-Bourboulis et al., 2020), yet mechanistic insights are still missing. Exacerbated immune responses play a major role in the pathophysiology of SARS, leading to severe lung injury and respiratory failure (Perlman and Dandekar, 2005). Mitigation of immunodysregulation could thus represent a major therapeutic avenue for the treatment and prevention of severe COVID-19 (Dimopoulos et al., 2020; Jamilloux et al., 2020). An early report investigating transcriptional profiles of peripheral blood mononuclear cells (PBMC) of 7 patients with mixed clinical courses of COVID-19, revealed complex immune deviations with changes in numerous cellular compartments, including monocytes, NK cells, dendritic cells and T cells (Salomé and Mahmood, 2020; Wilk et al., 2020).

The heterogeneity of clinical phenotypes and the complexity of systemic immune responses to COVID-19 highlight the need for detailed insights into different stages of the disease using high-resolution techniques and well-characterized clinical cohorts. Here, we hypothesized that distinct immune responses, particularly within the innate immune cell compartment, underlie the different clinical trajectories of COVID-19 patients (McKechnie and Blish, 2020). The striking alterations in cell counts and activation states of different innate immune cells in COVID-19 patients, and their relation to disease severity are currently not sufficiently understood. Here, we performed single-cell transcriptomics and single-cell proteomics on blood samples from two independent cohorts of COVID-19 patients, which allowed for instant cross-validation of immunological findings. COVID-19 patients with mild disease courses in both cohorts showed increased CD14^+^HLA-DR^hi^CD11c^hi^ inflammatory monocytes, which were marked by a strong IFN-stimulated gene (ISG^+^) signature. In contrast, severe COVID-19 was associated with the appearance of immature CD14^+^*MPO*^+^*Ki67*^+^*HLA-DR*^lo^ ISG^+^ suppressive monocytes and dysfunctional low-density neutrophils. The latter were identified as *ARG1^+^MPO^+^BPI+* pre-neutrophils and *ARG1^+^CD101^+^S100A8/A9^+^* immature neutrophils, indicative of emergency myelopoiesis. In addition, suppressive *PD-L1^+^CD123^+^* neutrophils were detected at late stages in severe COVID-19. Collectively, our study links highly dysregulated myeloid cell responses to severe disease courses of COVID-19.

## Results

### Dual center cohort study to assess immunological alterations in COVID-19 patients

In order to gain detailed insights into the distinct immunological events in mild *versus* severe COVID-19, we analyzed peripheral blood samples collected from independent cohorts of COVID-19 patients at two university medical centers in Germany. Samples from the Berlin cohort (cohort 1) were analyzed by mass cytometry (CyTOF) and single-cell RNA-sequencing (scRNA-seq) using a droplet-based single cell platform (10x Chromium), while samples from the Bonn cohort (cohort 2) were analyzed by multi-colour flow cytometry (MCFC) and scRNA-seq using a microwell-based system (BD Rhapsody). We profiled a total of 8.1 million cells by their surface protein markers and over 210,000 cells by scRNA-seq in 131 samples derived from a total of 33

### COVID-19 patients and 23 controls (Fig. 1A+B, S1A, Table S1)

We delineated COVID-19-induced alterations of the major leukocyte lineages by mass cytometry on whole blood samples from COVID-19 patients collected between day 6 and day 13 after disease symptom onset and compared them to age-matched healthy controls. Two antibody staining panels were designed to capture alterations in mononuclear cells (lymphocytes, monocytes and dendritic cells, panel 1), and in the granulocyte compartment (panel 2), respectively (**Table S2**). High-resolution SPADE analysis was performed with 400 target nodes and individual nodes were aggregated into cell subsets according to the expression of lineage-specific cell markers, such as CD14 for monocytes and CD15 for neutrophils (**Fig. S1B**). Visualization of t-distributed stochastic neighbor embedding (viSNE)-automated analysis revealed a clear separation of samples from COVID-19 patients and healthy controls, with marked changes of the monocyte- and the granulocyte compartment (**Fig. 1C**). Comparison of healthy control samples utilized for mass cytometry (HC CyTOF) to MCFC data from our recently published cohorts of healthy controls (HC flow) (Kverneland et al., 2016), demonstrated high similarities in the proportions of granulocytes, lymphocytes, T cells, total monocytes and CD14^lo^CD16^hi^ ‘non-classical’ monocytes in whole blood samples from healthy controls, irrespective of the applied methodology (**Fig. 1D**). This allowed us to use the absolute numbers of the published cohorts to also report differences in absolute numbers in COVID-19 samples. In line with recent reports (Barnes et al., 2020; Xintian et al., 2020), we observed a marked leukocytosis with increased proportions of granulocytes and neutrophils in patients with severe COVID-19 (**Fig. 1D**). In contrast, total lymphocyte- and T cell numbers were strongly reduced in all COVID-19 patients. Furthermore, we found a significant reduction of monocytes, particularly non-classical monocytes were virtually absent in COVID-19 (**Fig. 1D**). These changes, increased neutrophils in severe COVID-19 and loss of non-classical monocytes in both mild and severe disease, were validated in cohort 2 by MCFC (**Fig. S1C**).

**Figure 1.**
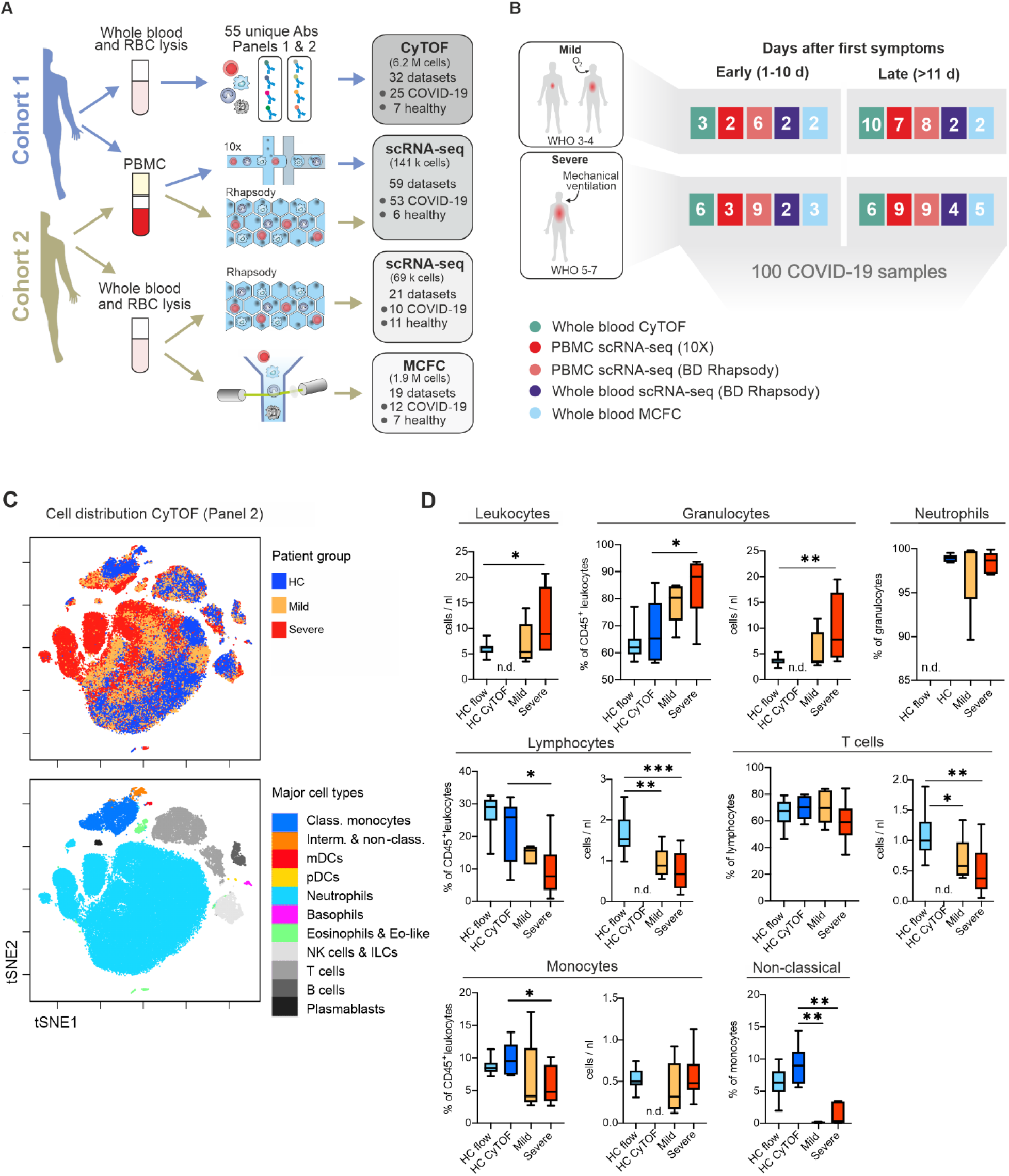
Cohort definition and single-cell multi-omics analysis strategy. **A**, Processing pipeline for healthy and COVID-19 blood samples strategies used in this study. PBMC are isolated after Ficoll, labeled with cell hashing antibodies and loaded on a droplet-based single-cell RNA-seq (scRNA-seq) platform. Red blood cells (RBC) are lysed and immune cells are labeled with two panels of metal-labeled antibodies and processed with CyTOF platform. Number of subjects analyzed for each cohort included in this study is summarized on the right boxes. **B**, Features of the cohort classified according to WHO-defined clinical grades (3 to 8) and the time after first symptoms. **C**, Visualization of t-distributed stochastic neighbor embedding (viSNE)-automated analysis of CD45+ leukocytes, down-sampled to 70,000 cells, from our mass cytometry analyses using antibody panel 2 based on similarities in expression of 29 markers as defined in supplementary table 2. Cells are colored according to donor origin (blue = healthy controls, yellow = mild COVID-19, red = severe COVID-19) and major lineage subtypes. **D**, Box plots summarising differences in major immune cell lineage subtype composition of whole blood samples from COVID-19 patients with mild (n=5) or severe disease (n=6) course, age-matched healthy controls utilized in mass cytometry (HC CyTOF, n=7) or measured by conventional or multi-colour flow cytometry (HC flow, n=18) as previously reported (Kverneland et al., 2016). *p<0.05, **p<0.01, p<0.001

Thus, SARS-CoV-2 infection induces lymphopenia and strong alterations within the myeloid compartment, with a drastic reduction of non-classical monocytes and neutrophilia in severe cases of COVID-19.

### Severity-dependent alterations of the myeloid cell compartment in COVID-19

Given the dramatic changes in various immune cell populations in COVID-19 (**Fig. 1C+D**), we next assessed their composition and activation status during the course of SARS-CoV-2 infection by droplet-based scRNA-seq in 21 samples from 12 COVID-19 patients (4 mild & 8 severe, cohort 1, **Table S1**) collected between day 7 and day 20 after disease symptom onset. A total of 25,667 single-cell transcriptomes of PBMC were analyzed together with 22,418 PBMC from a publicly available control dataset (4 healthy donors). Two-dimensional data representation using Uniform Manifold Approximation and Projection (UMAP) and high-resolution cell type classification identified all major cell types expected in the mononuclear compartment of blood, with a high granularity in the monocyte compartment as indicated by identification of four monocyte subsets (cluster 2, 5, 12, 14) (**Fig. 2A**). The top 10 genes identifying each cluster can be found in **Fig. S2A**. Monocytes in cluster 2, 5, and 14 expressed *CD14,* whereas cluster 12 comprised the non-classical monocytes marked by *FCGR3A* (CD16a). Separate visualization of cells in mild and severe cases, revealed highly disease severity-specific clusters (**Fig. 2B**). A distinct subset of the CD14^+^ monocytes marked by high expression of interferon (IFN)-stimulated genes (ISG, cluster 5), including *ISG15, IFI6, IFITM3,* or *APOBEC3A* (**Fig. 2C**), was selectively detected in mild COVID-19. The most prominent change in severe COVID-19, however, was the appearance of two cell populations (cluster 9+13, **Fig. 2B**), that were absent in PBMC of patients with mild COVID-19 and healthy donors. Based on published markers, the cell populations were identified as neutrophils and immature neutrophils (**Fig. 2A+B**). The immature neutrophils (cluster 13) expressed *CD24, MMP8, DEFA3* and *DEFA4,* whereas the identified neutrophil population (cluster 9) was marked by *FCGR3B* (CD16b), *CXCL8* (IL-8) and *CSF3R* (G-CSF receptor) expression (**Fig. 2C**). The fact that these neutrophil clusters migrated with PBMC fractions on a density gradient marked them as low-density neutrophils (LDN).

**Figure 2.**
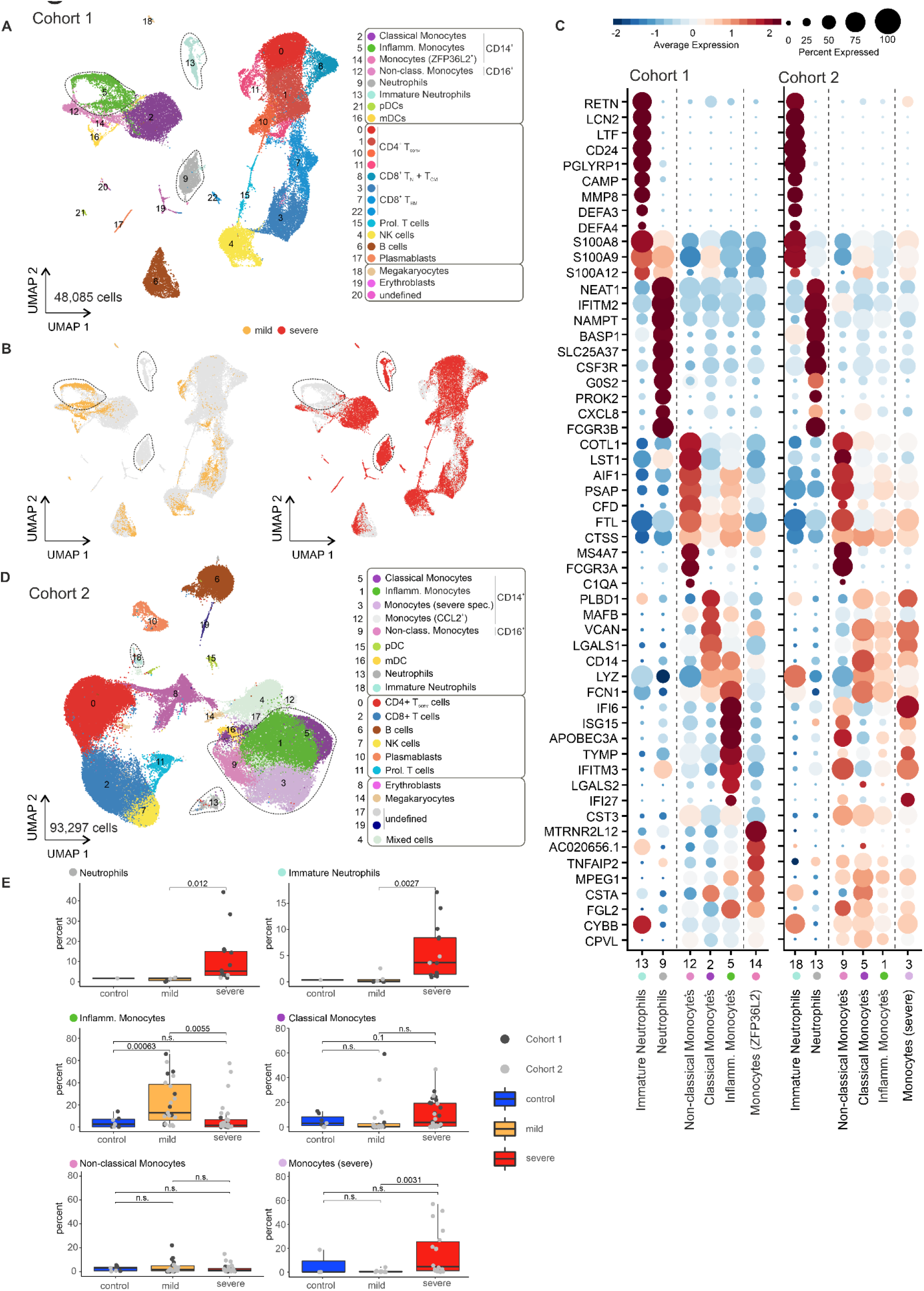
scRNA-seq of PBMC from patients of two independent cohorts reveals dynamic changes in the composition and the transcriptional states of myeloid cells in the peripheral blood in COVID-19. Abbreviations: HC: Healthy control. **A**, UMAP visualization of 10x chromium scRNA-seq profiles of 48.085 PBMC purified by density gradient centrifugation of peripheral blood from 21 samples of different time points of 4 mild and 8 severe patients in cohort 1 colored according to the indicated cell type classification based on Louvain clustering, reference-based cell-type annotation and marker gene expression patterns. **B**, UMAP shown in (a) colored according to disease severity (yellow = mild COVID-19, red = severe COVID-19). **C**, Dot plot representation of the top 10 marker genes sorted by average log fold change determined for the indicated myeloid cell subsets in the PBMC data set of cohort 1 plotted across the subsets of both cohorts. **D**, UMAP visualization of BD rhapsody scRNA-seq profiles of 93.297 PBMC purified by density gradient centrifugation from peripheral blood from 35 samples of 6 mild and 5 severe and 2 control patients of different time points in cohort 2 colored according to the indicated cell type classification based on Louvain clustering, reference-based cell-type annotation and marker gene expression patterns (Suppl. Fig. 2A). **E**, Box plots visualizing the percentages of the indicated cell subsets of the total number of PBMCs assessed per patient in the respective data set. Boxes are colored according to disease group and dots according to the respective cohort of the sample. For neutrophils and immature neutrophils only fresh PBMC samples were included.

In the second cohort, PBMC from 14 COVID-19 patients (7 mild, 7 severe) sampled between days 3 and 28 after onset of symptoms, and 2 controls, were collected for single-cell transcriptomic analysis using a microwell-based platform (BD Rhapsody). In total, high-quality single-cell transcriptomes for 93,297 PBMC were assessed and their overall population structure was visualized in two-dimensional space using UMAP (**Fig. 2D**). Data-driven cell type classification based on public reference transcriptome data (Aran et al., 2019) and cluster-specific marker gene expression depicted the presence of all major cell types expected in the PBMC compartment and revealed the appearance of additional clusters and substructures. The top 10 genes specifically expressed in each cluster of the UMAP depicted in **Fig. 2D** are shown in **Fig. S2B**. Similar to cohort 1, samples in cohort 2 also exhibited a prominent plasticity of the monocyte compartment, which could be subclassified into six clusters (**Fig. 2D**, cluster 1, 3, 5, 9, 12). Disease severity-associated changes seen in cohort 1 were also evident in cohort 2 (**Fig. 2E**). The appearance of LDN populations within the isolated PBMC fraction was validated in cohort 2, albeit at lower frequencies. Immature neutrophils and mature neutrophil cell clusters were detected in both cohorts (cluster 13 and 9 in cohort 1, cluster 18 and 13 in cohort 2) and showed a nearly identical marker gene expression profile (**Fig. 2C**). Similar to cohort 1, a prominent shift in subpopulation occupancy was observed in the monocyte clusters, particularly in cluster 3, 5, and 9 (**Fig. 2D+E**). Comparison of the specific marker genes identified for the monocyte-associated subcluster in cohort 1 with those in the corresponding cluster in cohort 2 revealed presence of the inflammatory ISG signature within the monocyte compartment, although it spread across different clusters in cohort 2 (cluster 9, 3, 1, **Fig. 2C**).

Single-cell transcriptomics of PBMC from COVID-19 patients identified disease-severity dependent cell subsets in the monocyte compartment as well as the appearance of two LDN populations.

### Mild COVID-19 is characterized by HLA-DR^+^ CD11c^+^ monocytes with an IFN signature

The monocyte compartment is particularly affected by COVID-19, as seen by mass cytometry revealing a loss of CD14^lo^CD16^hi^ non-classical monocytes, particularly in mild COVID-19 (**Fig. 1C+D**). Substantial shifts in monocyte subpopulation structure were also evident by scRNA-seq depending on disease severity (**Fig. 2**). To further dissect phenotypic alterations of the monocyte compartment in SARS-CoV-2 infection, we applied mass cytometry using a panel of 38 antibodies (**Table S2**, panel 1) to whole blood samples from COVID-19 patients with a mild or severe disease course, and corresponding age- and gender-matched healthy controls. Unsupervised cluster analysis using 15 surface antigens as well as the proliferation marker Ki67 separated the myeloid compartment into subpopulations of ‘classical’, ‘intermediate’ and ‘non-classical’ monocytes, myeloid dendritic cells (mDC) and plasmacytoid dendritic cells (pDC) (**Fig. 3A**). The classical monocyte compartment displayed a high level of heterogeneity and separated into two main subclusters. The majority of classical monocytes showed high expression of activation markers CD38, CD95, and CXCR3. Classical monocyte separated into two main subclusters based on high expression of CD62L and Ki67, indicative of proliferative capacity, *versus* high expression of CD226, CD69, HLA-DR and CD11c identifying highly activated, inflammatory cells (**Fig. 3A**). Monocytes from COVID-19 patients were clearly separated from those of healthy controls by viSNE analyses (**Fig. 3B**), mainly based on higher CD226 and CXCR3 expression in COVID-19 (**Fig. S3A**). Enhanced CD226 expression on activated monocytes might promote diapedesis through endothelial junctions and tissue infiltration, potentially explaining the relative reduction in overall monocytes in COVID-19 patients (Reymond et al., 2004). Classical monocytes in mild COVID-19 were enriched in HLA-DR^hi^ and CD11c^hi^ populations compared to severe disease and healthy controls (**Fig. S3A**). Thus, the response pattern of peripheral classical monocytes to SARS-CoV-2 infection is associated with disease severity.

**Figure 3.**
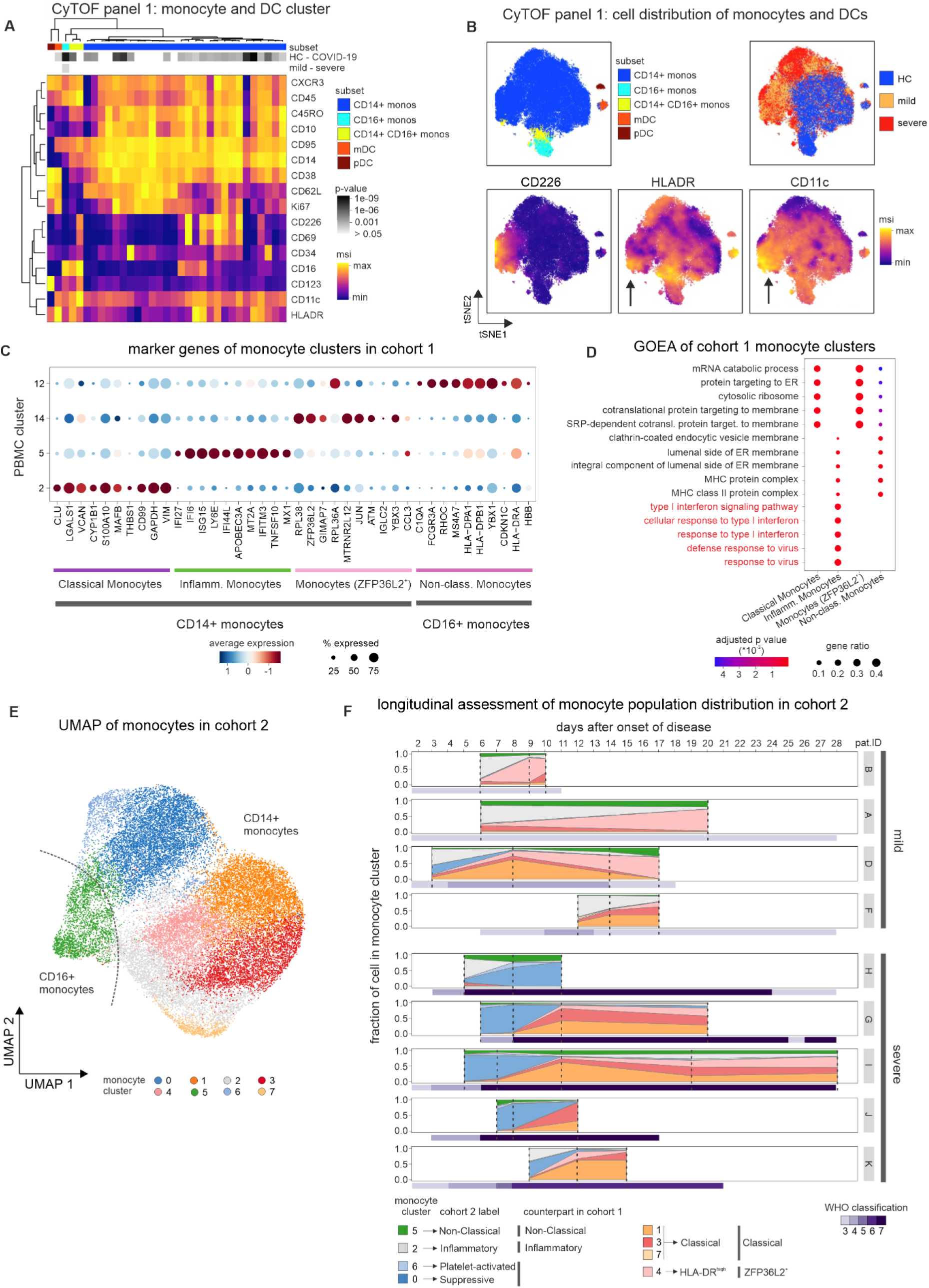
Subjects with different COVID-19 stages display unique phenotypical and transcriptional monocyte signatures. **A**, Heatmap revealing differences in marker expression determined by mass cytometry (CyTOF) using antibody panel 1 of monocyte and dendritic cell cluster. Cell clusters are displayed in columns and marker identity is indicated in rows. MSI = marker staining intensity respective expression level, significance level for the comparisons i) healthy controls (HC, n=7) versus COVID-19 (upper row) as well as ii) mild (n=5) versus severe (n=6, lower row) are indicated using a grey scale on top of the heatmap. **B**, t-SNE plots of monocytes and dendritic cells, down-sampled to 70,000 cells, based on their similarities in expression of 35 markers as defined in supplementary table 2. Cells are colored according to parental subclusters (classical monocytes, non-classical monocytes, intermediate monocytes, myeloid dendritic cells, plasmacytoid dendritic cells), donor origin (blue = healthy controls, yellow = mild COVID-19, red = severe COVID-19) and expression intensity of CD226, HLA-DR and CD11c. **C**, Dot plot representation of marker genes calculated for the clusters within the monocyte space of cohort 1. **D**, Gene ontology enrichment analysis of complete marker genes obtained for each identified monocyte population of cohort 1. **E**, UMAP visualization of 29,334 cells within the monocyte space of cohort 2. Cells are colored according to the identified clusters. **F**, Cluster occupancy per patient over time. Coloring according to the clusters identified in Figure 3E. Vertical dashed lines indicate the time points at which the samples were taken. The violet bar below the individual patient data indicates the respective WHO classification. Patient IDs are shown on the right.

To investigate molecular responses of monocytes to SARS-CoV-2 infection, we extracted clusters 2, 5, 12 and 14 from the PBMC dataset of cohort 1 (**Fig. 2A**) for further in-depth analysis. Monocytes in cluster 2 particularly expressed genes found in classical monocytes, such as *VCAN* and *S100A10,* while cells in cluster 14 were characterized by high expression of *ZFP36L2*, and were hence labeled as *ZFP36L2^+^* monocytes and classical monocytes, respectively (**Fig. 3C**). Cluster 12 was defined as non-classical monocytes by high expression of *FGR3A,* encoding CD16. As outlined before, the remaining cluster 5 (inflammatory monocytes) was characterized by an ISG program, which was further corroborated by gene ontology enrichment analysis (GOEA), assigning this cluster to ‘type I interferon signaling pathway’ (**Fig. 3D**). In addition, inflammatory (cluster 5) and non-classical monocytes (cluster 12) were also enriched for ‘MHC class II protein complex’ (MHCII), which is concordant with the high expression of *HLA-DRA* and *HLA-DRB1* in these cells (**Fig. 3C, S3B**). Expression of *CD14* indicated that the ISG^+^ cells represent a population of classical monocytes (**Fig. S3B**). Investigation of PBMC revealed that the number of these inflammatory monocytes differed between patients with mild and severe disease courses (**Fig. 2E**). We next investigated this shift in greater detail in cohort 2. The dimensionality reduction and re-clustering of the monocyte space revealed high transcriptional heterogeneity within the monocyte compartment (**Fig. 3E, S3D**). We identified three classical monocyte clusters (cluster 0, 2, and 6), which showed high ISG expression (**Fig. S3C+E**). Interestingly, only cluster 2 cells co-expressed high levels of MHCII molecules (**Fig. S3C**) and were thereby identified as counterparts of the inflammatory monocyte cluster in cohort 1 (**Fig. 3C**). In contrast, the remaining two ISG^+^ clusters exhibited low levels of *HLA-DRA* and *HLA-DRB1* expression (**Fig. S3D**), and cluster 6 was additionally characterized by platelet-associated genes *(TUBB1, PPBP,* and *PF4)* (**Fig. S3E**), indicating that these cells represent platelet-interacting inflammatory monocytes. Intriguingly, a proportion of cluster 0 monocytes expressed pre-maturation markers like *MPO* and *PLAC8* and *IL1R2,* which were recently linked to an immature monocyte population found in sepsis patients (Reyes et al., 2020) (**Fig. S3F**). Indeed, the gene signatures derived from these sepsis-associated monocytes were specifically enriched in clusters 0 and 6 (**Fig. S3G**). Low HLA-DR expression and monocyte immaturity result in reduced responsiveness to microbial stimuli (Veglia et al., 2018), which is why cluster 0 cells are referred to as suppressive monocytes. To understand how the identified cell populations in the patients change over time, we next investigated the time-dependent cluster occupancies per patient in cohort 2 (**Fig. 3F**). This analysis clearly showed that the ISG^+^ inflammatory clusters appeared in the early phase of the disease and gradually decreased over time. In addition, the inflammatory cluster 2 was associated with patients exhibiting a mild course of the disease, whereas the suppressive cluster 0 and platelet-activated monocyte cluster 6 were associated with severe disease. The gradual decrease of the ISG^+^ clusters was also evident in cohort 1, which showed a clear time-dependent decrease of *IFI6* and *ISG15* (**Fig. S3H**).

Taken together, we observed dynamic changes of the monocyte compartment in SARS-CoV-2-infected patients, associated with disease severity and time after onset of disease.

### Low-density neutrophils emerge in severe COVID-19 patients indicative of emergency myelopoiesis

Surprisingly, PBMC preparations in both cohorts contained two distinct clusters (**Fig. 2A**, clusters 9, 13; **Fig. 2D**, clusters 13, 18) of LDN, specifically in patients with severe disease. Since LDN in cohort 1 were more frequent than in cohort 2, we focused our in-depth analysis on cohort 1 data. We subsampled all LDN (**Fig. 4A**) and re-clustered the cells which gave rise to 8 transcriptionally distinct clusters within the LDN compartment of PBMC (**Fig. 4A+B**). We first analyzed previously described markers for pre-neutrophils, immature and mature neutrophils (Ng et al., 2019; Scapini et al., 2016) and identified cluster 4 as CD15*(FUT4)^+^CD63^+^*CD66b*(CEACAM8)^+^* pre-neutrophils, clusters 3 and 6 as CD11b*(ITGAM)^+^CD101^+^* immature neutrophils and the remaining clusters as mature neutrophils (**Fig. S4A**). The identification of pre-neutrophils and immature neutrophils was further supported by signature enrichment of neutrophil progenitors derived from previous single-cell data (**Fig. 4C**) (Pellin et al., 2019; Popescu et al., 2019). Cell cycle gene analysis further corroborated cluster 4 being pre-neutrophils with the highest proportion of cells per cluster showing a proliferative signature (**Fig. S4B**). Clusters 0, 1, 2 and 5 (originally from cluster 9 in **Fig. 2A**) were characterized by expression of the mature neutrophil markers *FCGR3A* (CD16) and *MME* (CD10) (**Fig. S4A**).

**Figure 4.**
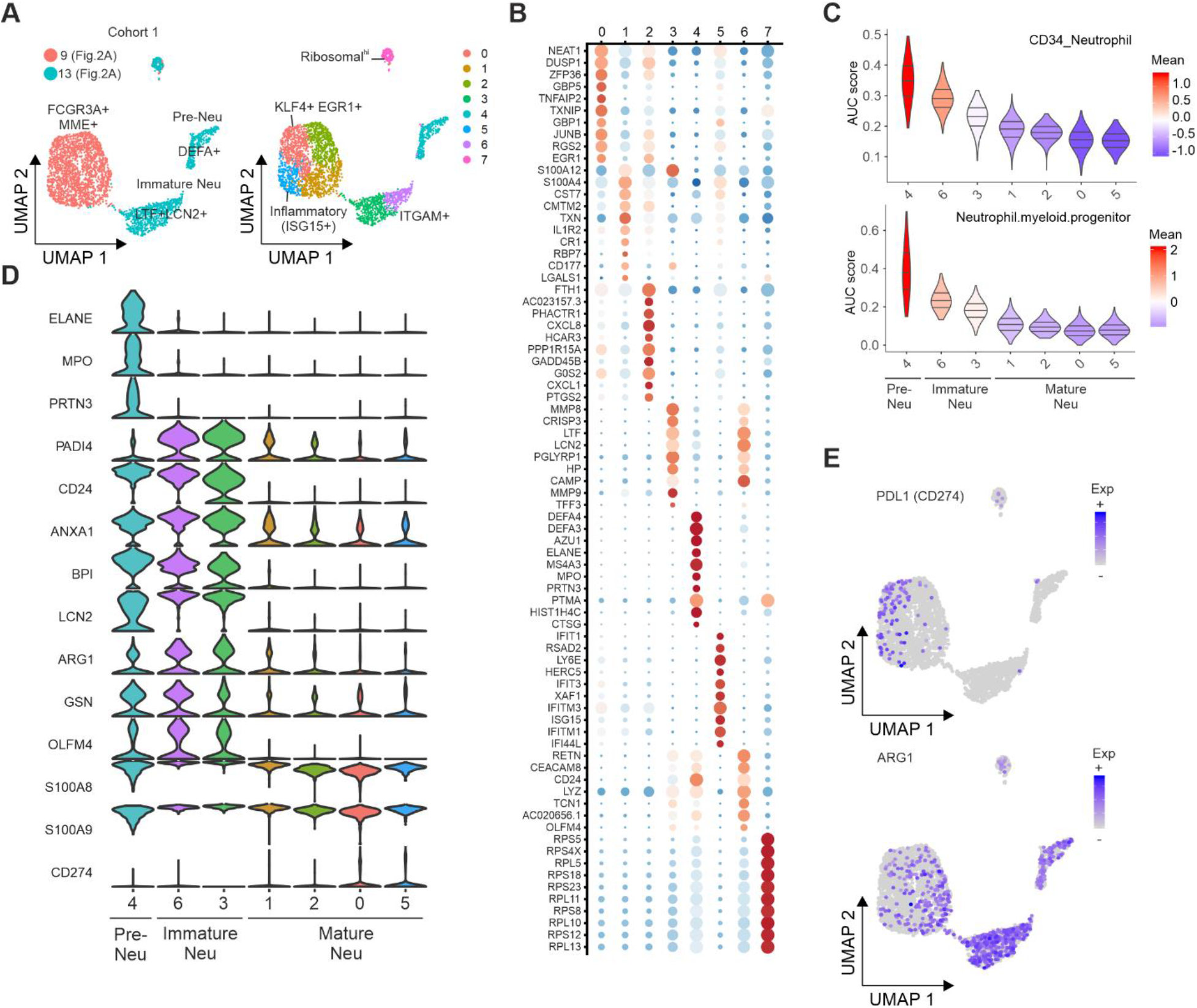
Immature and dysfunctional low-density neutrophils emerge in PBMC fractions. **A**, UMAP representation and clustering of the neutrophils identified in PBMC from cohort 1 (clusters 9 and 13 in Figure 2A). **B**, Dot plot visualization of the expression of the cluster-specific marker genes associated to each of clusters identified in panel A. **C**, Signature enrichment score of neutrophil progenitors derived from previous single-cell data. AUC: Area under the curve. **D**, Violin plots giving the expression of selected activation genes across the neutrophil clusters identified in panel A. **E**, Expression of *ARG1* and *PD-L1* projected on the UMAP from panel A.

Visualizing the most differentially expressed genes for each cluster revealed extensive phenotypic heterogeneity within the LDN compartment (**Fig. 4B**). For example, cluster 5 (ISG^+^ neutrophils) within the mature neutrophils highly expressed many ISGs *(ISG15, IFITM1/3* and *RSAD2),* strongly supporting an activated inflammatory neutrophil phenotype. Cluster 4 (pre-neutrophils) expressed several genes (e.g. *MPO, ELANE, PRTN3*) that have been associated with pathophysiological conditions including sepsis (Ahmad et al., 2019; Carbon et al., 2019; Silvestre-Roig et al., 2019). Similarly, immature neutrophils (clusters 3 and 6) expressed other genes (e.g. *CD24, LCN2)* previously associated with unfavorable outcome in sepsis (Kangelaris et al., 2015).

LDN mainly arise under pathological conditions, notably in infection and sepsis in the context of emergency myelopoiesis, and they have been associated with dysfunctional immune responses marked by combined immunosuppression and inflammation (Silvestre-Roig et al., 2019). We therefore investigated the expression of prominent genes previously linked to such pathological conditions (**Fig. 4D+E**). Pre-neutrophils strongly expressed *PRTN3, ELANE,* and *MPO,* genes that are involved in neutrophil extracellular trap formation (Stiel et al., 2018; Thomas et al., 2014; You et al., 2019) among other functions. Both pre-neutrophils and immature neutrophils expressed PADI4, another co-factor in NETosis (Leshner et al., 2012). NETs have recently been implicated in the pathogenesis of COVID-19 (Barnes et al., 2020; Zuo et al., 2020). Other genes, including *CD24, Olfactomedin (OLFM4), Lipocalin 2 (LCN2),* Bactericidal/permeability-increasing protein (*BPI*), previously associated with poor outcome in sepsis, were highly expressed in pre- and immature neutrophils (Kangelaris et al., 2015). We also observed a very strong expression of the alarmins *S100A8* and *S100A9,* which is not restricted to the pre-and immature state (**Fig. 4D**). Other members of the S100 gene family (e.g. *S100A4, S100A12*) are also strongly induced in different neutrophil clusters. Finally, known inhibitors of T cell activation, namely *PD-L1 (CD274)* and *Arginase 1 (ARG1)* (Bronte et al., 2003; Li et al., 2018) were highly expressed in neutrophils in COVID-19 patients (**Fig. 4E**). *PD-L1^+^* mature neutrophils (clusters 0, 5) resemble cells found in HIV-1 infected patients (Bowers et al., 2014). *ARG1+* cells were found among mature and immature neutrophils (clusters 1, 2, 3, 4 and 6) and did not overlap with *PD-L1* expression suggesting an emergence of different populations of suppressive neutrophils in COVID-19. *ARG1^+^* neutrophils in sepsis patients were shown to deplete arginine and constrain T cell function in septic shock (Darcy et al., 2014), and were predictive of the development of nosocomial infections (Uhel et al., 2017). Collectively, analyzing the LDN compartment recovered from PBMC fractions of COVID-19 patients revealed multiple mechanisms that might contribute to severe disease course and dysregulated immune responses.

### Persistent increase in activated and immunosuppressive neutrophils in severe COVID-19

To further dissect the profound alterations within the neutrophil compartment, we applied mass cytometry to whole blood samples from cohort 1 and age- and gender-matched healthy controls. We specifically designed the antibody panel 2 to not only identify neutrophils but also discriminate between maturation stages and reveal phenotypical signs of activation, immunosuppressive properties or dysfunction. Unsupervised clustering analysis of all neutrophils acquired from healthy control and COVID-19 samples identified 7 major subsets (**Fig. 5A**). Whereas neutrophils belonging to cell subsets 2, 5 and 7 appeared quiescent, with low expression of activation markers like CD64, neutrophil subsets 1 and 6 appeared to be dominated by highly activated, CD64^+^, Siglec 8^+^, RANK^+^ and RANKL^+^ as well as Ki67^+^ proliferating cells. Cells within subset 3 adopted an intermediate phenotype. Neutrophils from COVID-19 patients clearly separated from those of healthy controls, and neutrophils in patients with severe COVID-19 were distinct from those of patients with mild disease (**Fig. 5B**). Subset 1 was highly enriched for neutrophils from COVID-19 patients. Consequently, we observed an increase in CD34^+^ immature and activated neutrophils with high CD64, Siglec 8, RANK and RANKL (Riegel et al., 2012) expression in samples from COVID-19 patients regardless of disease course (**Fig. 5B+C**). In contrast, CD62L was exclusively downregulated in up to 50% of the neutrophils from severe but not mild COVID-19 patients, indicative of distinct dysfunctional neutrophil populations with immunosuppressive properties (Kamp et al., 2012; Pillay et al., 2012; Tak et al., 2017). These findings were further supported by upregulation of CD123 and PD-L1 expression (**Fig. 5B+C**) as hallmarks of myeloid-derived suppressor cell (MDSC) function (Cassetta et al., 2019; Testa et al., 2004; Younos et al., 2015) on neutrophils of severe COVID-19 patients. In addition, neutrophils of severe COVID-19 patients adopted an aged phenotype with elevated CD45 expression (**Fig. 5B**).

**Figure 5.**
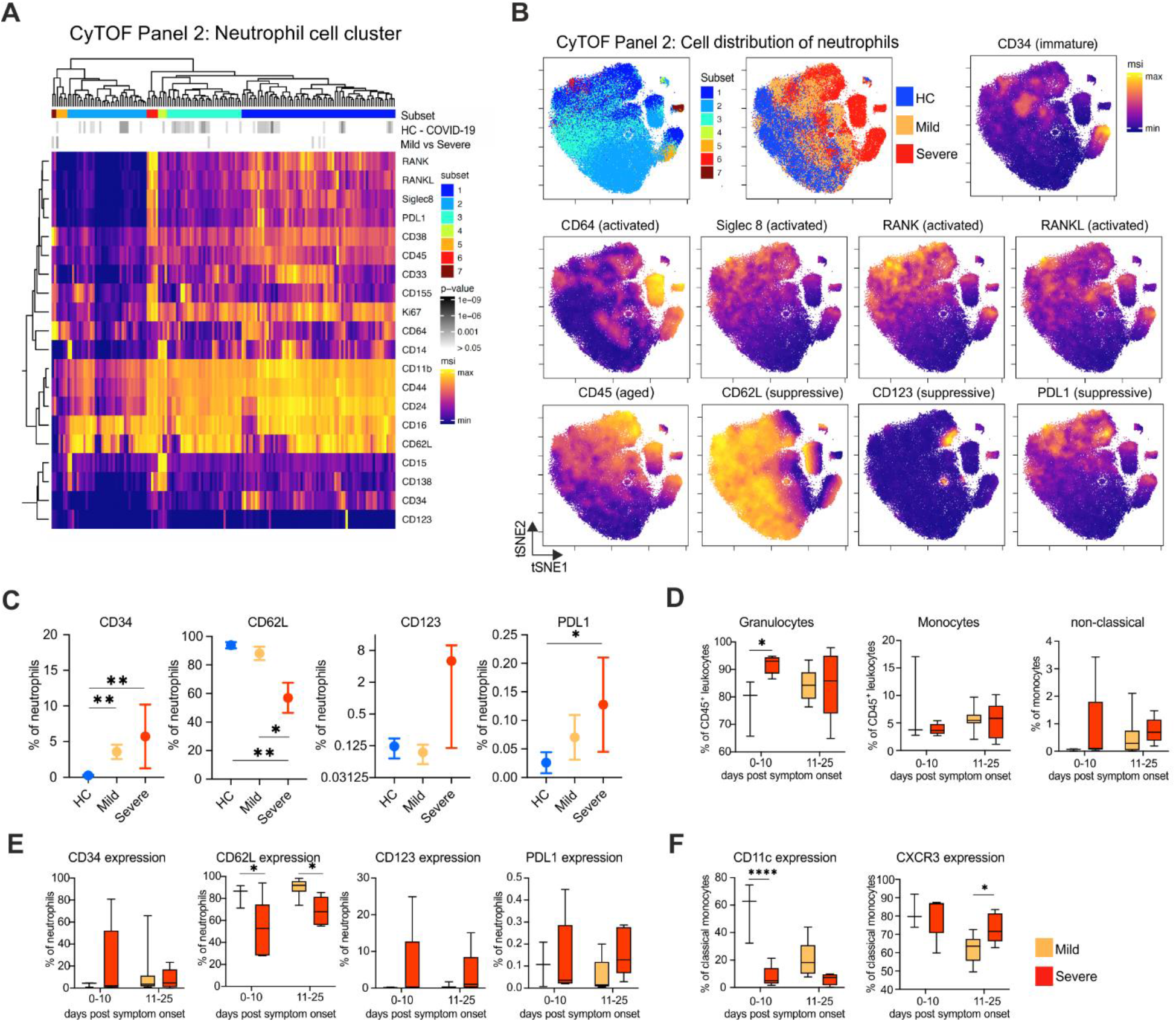
Disbalance between increase in activated and potentially immunosuppressive neutrophils discriminates between mild and severe COVID-19 patients. **A**, Heatmap revealing differences in marker expression determined by mass cytometry (CyTOF) using antibody panel 2 of identified neutrophil cell subcluster (1 to 7). Neutrophil cell clusters belonging to the identified subcluster are displayed in columns and marker identity is indicated in rows. MSI = marker staining intensity respective expression level, significance level for the comparisons i) healthy controls (HC) versus COVID-19 (upper row) as well as ii) mild versus severe (lower row) are indicated using a grey scale on top of the heatmap. **B**, t-SNE plots of neutrophils, down-sampled to 70.000 cells, based on their similarities in expression of 29 markers as defined in supplementary table 2. Cells are colored according to neutrophil subcluster, donor origin (blue = healthy controls, yellow = mild COVID-19, red = severe COVID-19) and expression intensity of CD34, CD64, Siglec 8, RANK, RANKL, CD45, CD62L, CD123 and PD-L1. **C**, Bar graphs summarizing CD34, CD62L, CD123 and PD-L1 expression as % positive cells within neutrophils of whole blood samples from COVID-19 patients with mild (n=5) or severe disease (n=5) course and age-matched healthy controls (n=7). *p<0.05, **p<0.01 **D-F**, Box plots summarizing time-dependent differences in total granulocytes and monocytes, non-classical monocytes (D) CD34, CD62L, CD123 and PD-L1 expressing neutrophils (**E**) as well as CD11c and CXCR3 expressing classical monocytes (**F**) measured by mass cytometry in whole blood samples from cohort 1 distinguishing between COVID-19 patients with mild (days 0-10: n=3, days 11-25: n=10) or severe/critical disease (days 0-10: n=5, days 11-25: n=5) course. *p<0.05, *p<0.0001

Thus, SARS-CoV-2 infection induces major alterations in the neutrophil compartment. While neutrophils in patients with mild COVID-19 display an activated phenotype, additional markers of immunosuppression or dysfunction are upregulated in patients with severe disease.

The differences in the functional states of neutrophils in mild and severe COVID-19 in the snap-shot analysis, prompted us to perform a longitudinal analysis. We analyzed samples according to collection either within the first 10 days (early) or during the second and third week (late) after onset of disease symptoms. The proportional increase in total granulocytes and CD34^+^ neutrophil subset in severe COVID-19 was only detected during the early phase of the disease (**Fig. 5D+E**). However, the additionally observed alterations in the neutrophil compartment such as loss of CD62L expression accompanied by increase of CD123 and PD-L1 positive populations as signs of dysfunctionality and immunosuppressive properties were persistent in severe COVID-19 patients (**Fig. 5E**).

Next, we analyzed whether similar to the evolution on the transcriptional level, we can also capture time-dependent changes of the monocyte compartment on the protein level. The proportion of non-classical monocytes started to recover in mild patients during the later stages of the disease (**Fig. 5D**). This was accompanied by reduced CD11c and CXCR3 expression in classical monocytes (**Fig. 5F**) and relates very well to the described longitudinal changes of the HLA-DR & ISG^+^ monocyte cluster (**Fig. 3F, S3H**). In contrast, although classical monocytes of severe patients never showed high CD11c expression, they maintained high levels of CXCR3 expression even in late stages of the disease, indicating prolonged activation (**Fig. 5F**). Thus, severe COVID-19 patients are characterized by a combination of persistent inflammation and immunosuppression, which is reminiscent of long-term post-traumatic complications (Hesselink et al., 2019). Our data reveal an early strong monocyte activation versus persistent signs of neutrophil dysfunctionality as discriminators between mild and severe COVID-19.

### Single cell transcriptomes of whole blood neutrophils reveal suppressive signatures in severe COVID-19

Whole blood CyTOF analysis (cohort 1) clearly indicated a very distinct molecular regulation within the neutrophil compartment in severe and mild COVID-19. To further delineate this regulation with respect to the underlying transcriptional programs, we performed scRNA-seq analysis on fresh whole blood samples from 19 individuals (21 samples, cohort 2). Overlay of all samples of cohort 2 (fresh/frozen PBMC, fresh whole blood, 162,697 cells, **Fig. S6A**), without batch correction, revealed the major cell type distribution, including the granulocyte compartment (**Fig. 6A, S6A**). Cell type distribution as identified by scRNA-seq (**Fig. S6B**) strongly correlated with the MCFC data of the same samples (**Fig. S6C**). We noticed abnormal transcriptional activation in six samples, which was caused by protracted cold treatment (4°C) prior to single-cell sampling, as recently described for PBMC (Massoni-Badosa et al., 2020). These samples were therefore excluded from further analysis. The remaining 30,019 neutrophils were subsampled and revealed a structure of 11 clusters (**Fig. 6B-C**). Using marker- and data-driven approaches as applied to LDN (**Fig. 4D, S4A**), we identified CD15*(FUT4)^+^CD63^+^CD66b^+^CD101^+^CD10^-^* pre-neutrophils, CD11b*(ITGAM)^+^CD101^+^CD10^-^* immature neutrophils along with 9 mature neutrophil clusters (**Fig. 6B-D, S6D-E**). Heterogeneous expression of various markers involved in neutrophil function including *SELL* (L-selectin, CD62L), *CXCR2,* FCGR2 (CD32), and *CD10* as a marker for mature neutrophils, pointed towards distinct functionalities within the neutrophil compartment (**Fig. 6C, 6H, S6E**). The phenotypic diversity was further corroborated, when we assessed the expression of markers previously identified by CyTOF to be differentially regulated in patients with severe COVID-19 (**Fig. 5**). For example, we found elevated expression of *CD274* (PD-L1) and *CD64* in neutrophil clusters identified specifically in patients with severe COVID-19 (**Fig. 6E, S6F**). Out of the 11 neutrophil clusters identified in whole blood in cohort 2, 10 clusters could also be mapped to fresh PBMC samples in cohort 1 (**Fig. S6D**), indicating that scRNA-seq of fresh PBMC - in COVID-19 patients - reveals relevant parts of the neutrophil compartment. When we analyzed PBMC samples of cohort 2, these clusters could be identified in fresh samples, but were lost upon freezing (**Fig. S6A**). Importantly, the analysis corroborated the transcriptional phenotype of pre-neutrophils and immature neutrophils (cluster 8+9) in cohort 2 (**Fig. 6B-D, S6D-E**).

**Figure 6.**
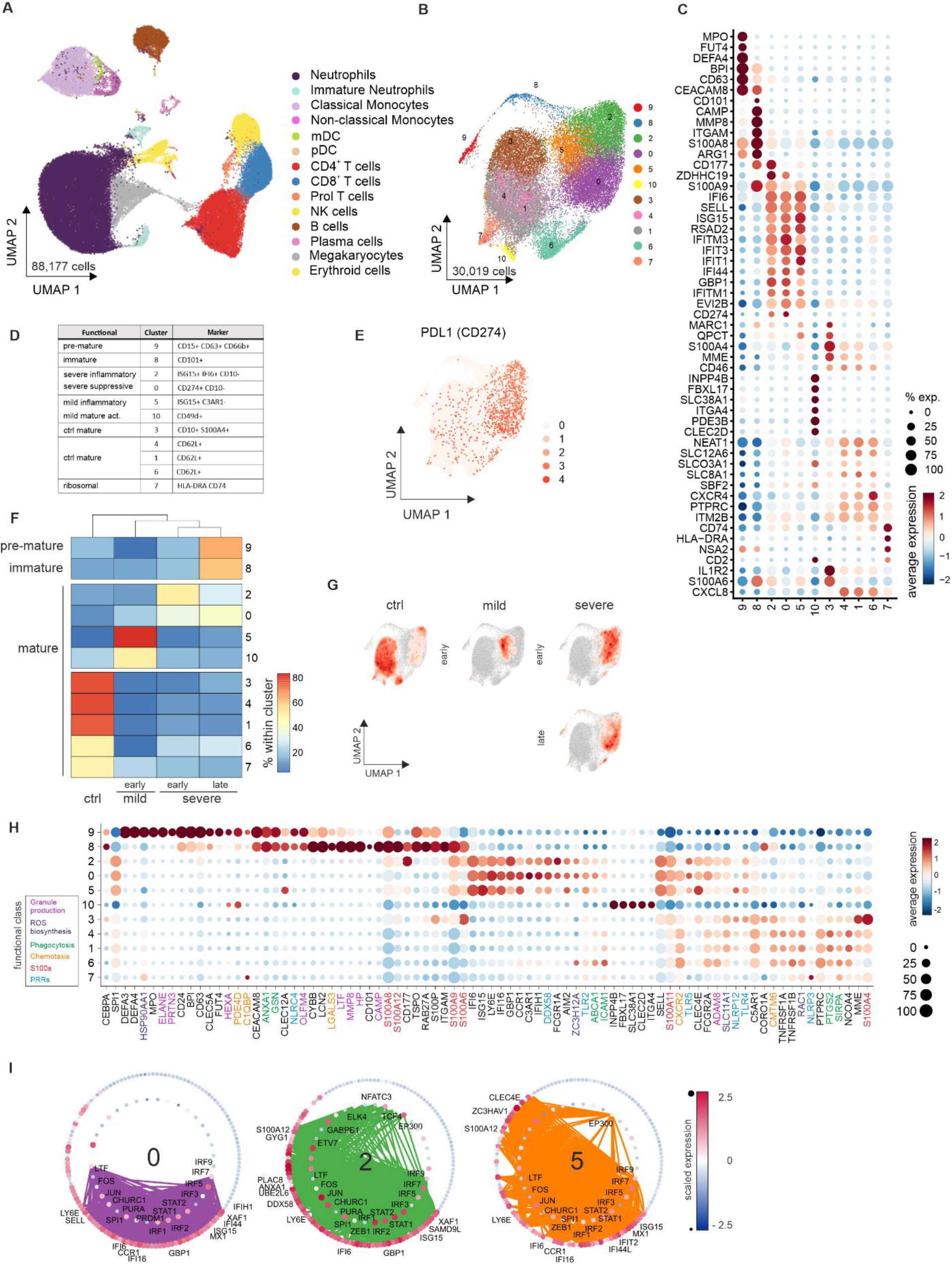
Delineation of suppressive mature neutrophils in late stage severe COVID-19. **A**, UMAP visualization of fresh blood samples from cohort 2 (PBMC and whole blood combined, 88,177 cells, controls (ctrl = 12), COVID-19 mild (mild = 6), COVID-19 severe (severe = 8). **B**, UMAP visualization of the immature and mature neutrophils (30,019 cells) from Figure 6A after exclusion of one experimental batch (see Methods, controls (ctrl = 10), COVID-19 early mild = 1, COVID-19 severe (early = 2, late = 2)). **C**, Dot plot of selected top 6 marker genes ordered by average log fold-change highlighting the heterogeneity of the neutrophil space in b. **D**, Overview of functional nomenclature and marker genes for each cluster in the neutrophil space in COVID-19 and control patients. **E**, UMAP visualization of neutrophils showing the scaled expression of PD-L1 *(CD274)* with a clear enrichment in the Covid-19 specific clusters 2 and 0. **F**, Confusion matrix for each cluster in Figure 6B divided by disease severity and time point showing the enrichment of each patient group for each cluster; the scale is normalized for each cluster. **G**, Density plot of cell frequency by disease severity and time point overlaid on UMAP visualization of the neutrophil space. **H**, Dot plot of genes from different functional classes showing the differences of the neutrophil states (based on literature research). **I**, Transcription factor binding prediction results for clusters 0, 2 and 5 shown as networks of transcription factors and their targets among the specifically expressed genes for the given cluster. Edges represent predicted transcriptional regulation. Transcription factors in the inner circle and their predicted target genes in the outer circle are represented as nodes sized and colored according to the scaled expression level across all clusters. Top 10 highest connected transcription factors and those exclusively predicted for the cluster as well as the top 5 highest connected target genes together with a literature-based selection of targets are labeled.

Heatmap and UMAP visualization of the cell type distribution identified pre-neutrophils and immature neutrophils in severe COVID-19, at late time points (**Fig. 6F-G**). Furthermore, clusters 2 and 0 were associated with severe COVID-19, while clusters 5 and 10 were found in mild COVID-19 (**Fig. 6F-G**). IFN-signatures were detected in neutrophils from mild and severe COVID-19 patients and even in some of the clusters derived from controls (**Fig. S6G**). However, expression of *CD177, CD274* (PD-L1) and zinc finger DHHC domain-containing palmitoyl transferases *ZDHHC19,* indicative of an immature and suppressive granulocyte phenotype (Volkmann et al., 2020), was specifically observed in patients with severe COVID-19 (**Fig. 6C**). ZDHHC19 has been shown to promote palmitoylation of STAT3 (Niu et al., 2019), as well as the Chikungunya virus derived protein nsP1 (Zhang et al., 2018). In addition, neutrophils in severe COVID-19 expressed *FCGR1A, ICAM1* and *ZC3H12A* (**Fig. 6H**), indicative of their suppressive nature. As indicated by the clustering in the UMAP (**Fig. 6B+G**), pseudotime analysis strongly suggested that cells in cluster 2 preceded those in cluster 0, and particularly *CD274* (PD-1L) was among the genes that further enriched in cluster 0 compared to cluster 2, indicating a progression towards a suppressive phenotype in late stage disease (**Fig. S6H+I**). Predictions of transcription factor (TF)-based regulation of the cluster-specific gene signatures revealed a strict separation of mature neutrophils from pre-neutrophils and immature neutrophils (**Fig. S6J**), but showed a surprisingly high overlap between clusters 5 (mild), 2 (severe early), and 0 (severe late) in STAT- and IRF-family member TFs (**Fig. 6I**). However, there was a striking loss of important TF hubs in cluster 0, found in late stage severe COVID-19 (**Fig. 6I**).

Collectively, the neutrophil compartment in peripheral blood of COVID-19 patients is characterized by the appearance of LDN, *ARG1^+^MPO^+^BPI^+^* pre-neutrophils and *CD10^+^ S100A8/A9^+^* immature neutrophils, reminiscent of emergency myelopoiesis, as well as *CD274*^+^(PD-L1^+^) suppressive mature neutrophils. All neutrophil subsets emerging in severe COVID-19 express an armamentarium of genes associated with suppressive functions as they have been described for sepsis or ARDS, indicating that a dysregulated myeloid cell compartment contributes to severe COVID-19.

## Discussion

SARS-CoV-2 infection generally causes mild disease in the majority of individuals, however about 10-20% of COVID-19 patients progress to severe disease with pneumonia and respiratory failure. The case-fatality ratio was found to be 49% among patients with critical illness and respiratory failure (Wu and McGoogan, 2020). The pathomechanisms that govern progression from mild COVID-19 to potentially fatal courses of disease are currently unknown, but dysregulated immune responses have been repeatedly described in patients with severe COVID-19 (Zhou et al., 2020c). Detailed knowledge of the underlying molecular processes is urgently needed in order to identify predictive biomarkers and therapeutic targets for severe COVID-19, which could enable individualized treatments.

Here, we employed five complementary technologies at single-cell resolution to assess differences in the systemic immune response in patients with mild or severe courses of COVID-19. We analyzed a total of 131 samples from two independent cohorts collected at two university medical centers in Germany. The combination of single-cell transcriptomics and single-cell proteomics, using different technological platforms and sample-processing strategies in independent patient cohorts provided unprecedented insights into the systemic immune responses in COVID-19 and allowed for cross-validation of key findings.

This multipronged approach revealed drastic changes within the myeloid cell compartment, particularly in patients with a severe course of disease. Early activation of HLA-DR^hi^CD11c^hi^ monocytes with a strong antiviral IFN-signature was a hallmark of mild COVID-19, which receded during the natural course of disease, as demonstrated at later time points in patients with mild COVID-19. In contrast, we found clear evidence of emergency myelopoiesis with release of pre-neutrophils and immature neutrophils with immunosuppressive features in severe COVID-19. Prolonged partial monocyte activation and release of dysfunctional neutrophils may thus contribute to severe disease course and ARDS development.

Several reports have described inflammatory monocyte responses, with a strong IFN-signature in COVID-19 (Liao et al., 2020; Merad and Martin, 2020; Zhou et al., 2020c). Mononuclear phagocytes and neutrophils appear to dominate inflammatory infiltrates in the lungs, and resident alveolar macrophages are replaced by inflammatory monocyte-derived macrophages in patients with severe COVID-19 (Liao et al., 2020). Here, we report substantial alterations of the monocyte compartment, with time- and disease severity-dependent changes in two separate cohorts of COVID-19 patients. We observed a depletion of CD14^lo^CD16^hi^ non-classical monocytes in all patients, which was particularly prominent in mild cases and less pronounced in severe disease (**Fig. 1D**). This is in line with previous reports, and it has also been observed in other severe viral infections (Lüdtke et al., 2016; Naranjo-Gómez et al., 2019). Strikingly, we found a strong transient increase in highly activated CD14^+^CD11c^hi^HLA-DR^hi^ ISG^+^ monocytes in mild cases of COVID-19, which were absent in severe cases (**Fig. 3**). In contrast, we identified immature suppressive monocytes with low expression of HLA-DR in patients with severe disease. Low HLA-DR expression on monocytes has been previously associated with an increased risk to develop infectious complications after trauma (Hoffmann et al., 2017) and increased mortality in septic shock (Monneret et al., 2006), Furthermore, CD14^+^HLA-DR^lo/neg^ monocytes, have also been identified as important mediators of tumor-induced immunosuppression (Mengos et al., 2019). Indeed, the severe COVID-19-specific immunosuppressive monocyte cluster showed enrichment of genes previously identified in sepsis patients (**Fig. S3H**) (Reyes et al., 2020).

Acute pathological insults, such as trauma or severe infections, trigger a process referred to as emergency myelopoiesis in order to replenish functional granulocytes and other hematopoietic cells. Emergency myelopoiesis is characterized by the mobilization of immature myeloid cells, which exert immunosuppressive functions (Loftus et al., 2018; Schultze et al., 2019). Emergence of suppressive myeloid cells has been previously observed during sepsis and severe influenza virus infection (Darcy et al., 2014; Loftus et al., 2018; Sander et al., 2010; De Santo et al., 2008). Here we detected pre-neutrophils and immature neutrophils within PBMC, indicating their reduced cellular density, specifically in severe COVID-19 (**Fig. 4**). These LDN showed an immature gene expression profile and a surface marker and gene expression profile indicative of immunosuppressive functions. For example, olfactomedin-4^+^ LDN have been associated with immunopathogenesis in sepsis (Alder et al., 2017).

Suppressive neutrophils have been shown to increase in patients with bacterial sepsis and their frequencies correlated with sepsis severity and systemic inflammation (Darcy et al., 2014), as well as in healthy volunteers treated with G-CSF (Hartung et al., 1995). Analyzing whole blood samples without density separation, we identified neutrophils with an IFN-signature in mild and severe COVID-19, but only in patients with severe disease, these activated neutrophils also expressed PD-L1 *(CD274),* which further increased in expression in late stage disease. The expression of *CD177* on mature activated neutrophils at early stages, but not at later stages, and the identification of genes associated with suppressive cellular functions *(ZDHHC19, ZC3H12A)* strongly favor a model in which neutrophils emerging prematurely from the bone marrow are programmed towards a suppressive program in severe COVID-19. The transcriptional programs induced in pre-neutrophils, immature and mature COVID-19-associated neutrophil clusters may also align with other clinical observations in severe COVID-19 patients, including increased NET formation (Barnes et al., 2020; Zuo et al., 2020), coagulation (Klok et al., 2020; Pfeiler et al., 2014) and immunothrombosis (Stiel et al., 2018; Xu et al., 2020). In contrast, in patients with mild COVID-19, none of these transcriptional programs was observed. Despite the fact that SARS-CoV2^neg^ controls exhibited a range of comorbidities (e.g. COPD, type diabetes II), we did not observe this massive expansion of suppressive neutrophils in these chronic inflammatory conditions. The pathophysiological consequences of this suppressive signature in severe COVID-19 are unclear at this stage, but it is highly likely that they contribute to immunoparalysis in critically ill patients, potentially leading to insufficient host defense, disbalanced inflammation and increased susceptibility to superinfections.

Collectively, our data link a striking appearance of immature and suppressive cells, in both the monocyte and neutrophil compartment, to disease severity in COVID-19. Consequently, for the development of better treatments and prevention of severe COVID-19, we may benefit from achievements in other fields such as cancer, which have successfully applied therapies targeting suppressive myeloid cells.

## STAR Methods

### Study subjects

#### Cohort 1 / Berlin cohort

This study includes a subset of patients enrolled between March 2 and March 27 2020 in the Pa-COVID-19 study, a prospective observational cohort study assessing pathophysiology and clinical characteristics of patients with COVID-19 at Charité Universitätsmedizin Berlin (Kurth et al., 2020). The study is approved by the Institutional Review board of Charité (EA2/066/20). Written informed consent was provided by all patients or legal representatives for participation in the study. The patient subset included in this analysis contains 7 healthy donors (**Table S1**) and 19 COVID-19 patients (**Figure 1A+B, Table S1**). All COVID-19 patients tested positive for SARS-CoV-2 RNA in nasopharyngeal swabs.

#### Cohort 2 / Bonn cohort

This study was approved by the Institutional Review board of the University Hospital Bonn (073/19 and 134/20). After providing written informed consent, 12 control donors (**Table S1**) and 14 COVID-19 patients (**Figure 1A+B, Table S1**) were included in the study. In patients who were not able to consent at the time of study enrollment, consent was obtained after recovery. COVID-19 patients who tested positive for SARS-CoV-2 RNA in nasopharyngeal swabs were recruited at the Medical Clinic I of the University Hospital Bonn between March 30 and May 17, 2020.

Isolation of blood cells for scRNA-seq

#### Cohort 1 / Berlin cohort

PBMC were isolated from heparinized whole blood by density centrifugation over Pancoll (density: 1.077g/ml; PAN-Biotech™). If the pellet was still slightly red, remaining CD235ab^+^ cells (Erythrocytes) were depleted by Negative Selection *(MagniSort™* Thermo Fisher). Subsequently the PBMC were prepared for 3’scRNA-seq (10xGenomics) or cryopreserved and analyzed later.

#### Cohort 2 / Bonn cohort

In the Bonn cohort, scRNA-seq was performed on fresh whole blood, fresh PBMC and frozen PBMC. Briefly, PBMC were isolated from EDTA-treated or heparinized peripheral blood by density centrifugation over Pancoll or Ficoll-Paque density centrifugation (density: 1.077g/ml). Cells were then washed with DPBS, directly prepared for scRNA-seq (BD Rhapsody) or cryopreserved in RPMI-1640 with 40% FBS and 10% DMSO. Whole blood was prepared by treatment of 1 ml peripheral blood with 10 ml of RBC lysis buffer (Biolegend). After one wash in DPBS cells were directly processed for scRNA-seq (BD Rhapsody) or MCFC. Frozen PCMC were recovered by rapidly thawing frozen cell suspensions in a 37° C water bath followed by immediate dilution in pre-warmed RPMI-1640+10% FBS (Gibco) and centrifugation at 300g for 5 min. After centrifugation, the cells were resuspended in RPMI-1640+10% FBS and processed for scRNA-seq. Antibody cocktails were cryopreserved as described before (Schulz et al., 2019).

### Antibodies used for mass cytometry

All anti-human antibodies pre-conjugated to metal isotopes were obtained from Fluidigm Corporation (San Francisco, US). All remaining antibodies were obtained from the indicated companies as purified antibodies and in-house conjugation was done using the MaxPar X8 labeling kit (Fluidigm). Supplementary table 2 shows a detailed list of all antibodies used for panel 1 and panel 2.

### Sample processing, antigen staining and data analysis of mass cytometry-based immune cell profiling

500 μl of whole blood (heparin) was fixed in 700 μl of proteomic stabilizer (Smart Tube Inc., San Carlos, US) as described in the user manual and stored at −80°C until further processing. Whole blood samples were thawed in Thaw/Lyse buffer (Smart Tube Inc.). For barcoding antibodies recognising human beta-2 microglobulin (B2M) were conjugated in house to ^104^Pd, ^106^Pd, ^108^Pd, ^110^Pd, ^198^Pt (Mei et al., 2015, 2016; Schulz and Mei, 2019). Up to 10 individual samples were stained using a staining buffer from Fluidigm with a combination of two different B2M antibodies for 30 min at 4°C. Cells were washed and pooled for surface and intracellular staining.

For surface staining the barcoded and pooled samples were equally divided into two samples. Cells were resuspended in antibody staining cocktails for panel 1 or panel 2 respectively (**Table S2**) and stained for 30 min at 4°C. For secondary antibody staining of panel 2, cells were washed and stained with anti-APC ^163^Dy for 30 min at 4°C. After surface staining cells were washed with PBS, resuspended in cell-ID intercalator ^103^Rh to discriminate between live and dead cells and incubated for 5 min at room temperature. After washing, cells were fixed overnight in 2 % PFA solution diluted dissolved in PBS to 2%.

For intracellular staining cells were washed twice with a permeabilization buffer (eBioscience, San Diego, US) and stained with the respective antibodies diluted in a permeabilization buffer for 30 min at room temperature. After washing, cells were stained with iridium intercalator (Fluidigm) diluted in 2 % PFA for 20 min at room temperature. Cells were washed once with PBS and then twice with ddH_2_O and kept at 4°C until mass cytometry measurement.

A minimum of 100.000 cells per sample and panel were acquired on a CyTOF2/Helios mass cytometer (Fluidigm). For normalization of the fcs files 1:10 EQ Four Element Calibration Beads (Fluidigm) were added. Cells were analyzed using a CyTOF2 upgraded to Helios specifications, with software version 6.7.1014, using a narrow bore injector. The instrument was tuned according to the manufacturer’s instructions with tuning solution (Fluidigm) and measurement of EQ four element calibration beads (Fluidigm) containing 140/142Ce, 151/153Eu, 165Ho and 175/176Lu served as a quality control for sensitivity and recovery. Directly prior to analysis, cells were resuspended in ddH_2_O, filtered through a 20-μm cell strainer (Celltrics, Sysmex), counted and adjusted to 5-8 × 105 cells/ml. EQ four element calibration beads were added at a final concentration of 1:10 v/v of the sample volume to be able to normalize the data to compensate for signal drift and day-to-day changes in instrument sensitivity. Samples were acquired with a flow rate of 300-400 events/s. The lower convolution threshold was set to 400, with noise reduction mode turned on and cell definition parameters set at event duration of 10-150 pushes (push = 13 μs). The resulting flow cytometry standard (FCS) files were normalized and randomized using the CyTOF software’s internal FCS-Processing module on the non-randomized (‘original’) data. The default settings in the software were used with time interval normalization (100 s/minimum of 50 beads) and passport version 2. Intervals with less than 50 beads per 100 s were excluded from the resulting FCS file.

### Blood processing for flow cytometry

1 ml of fresh blood from control or COVID-19 donors was treated with 10 ml of RBC lysis buffer (Biolegend). After RBC lysis, cells were washed with DPBS and 1-2 million cells were used for flow cytometry analysis. Cells were then stained for surface markers (**Table S3**) in DPBS with BD Horizon Brilliant™ Stain Buffer (Becton Dickinson) for 30 min at 4°C. To distinguish live from dead cells, the cells were incubated with LIVE/DEAD Fixable Yellow Dead Cell Stain Kit (1:1000 – Thermo Scientific). Following staining and washing, the cell suspension was fixed with 4% PFA for 5 min at room temperature to prevent any possible risk of contamination during acquisition of the samples. Flow cytometry analysis was performed on a BD Symphony instrument (Becton Dickinson) configured with 5 lasers (UV, violet, blue, yellow-green, red).

### 10x Genomics Chromium single-cell RNA-seq

PBMC were isolated and prepared as described above. Afterwards, patient samples were hashtagged with TotalSeq-A antibodies (Biolegend) according to the manufacturer’s protocol for TotalSeq^TM^-A antibodies and cell hashing with 10x Single Cell 3’ Reagent Kit v3.1. 50 μL cell suspension with 1×10^6^ cells were resuspended in staining buffer (2% BSA, Jackson Immuno Research; 0,01% Tween-20, Sigma-Aldrich; 1x DPBS, Gibco) and 5 μL Human TruStain FcX^TM^ FcBlocking (Biolegend) reagent were added. The blocking was performed for 10 min at 4°C. In the next step 1 μg unique TotalSeq-A antibody was added to each sample and incubated for 30 minutes at 4°C. After the incubation time 1.5 mL staining buffer were added and centrifuged for 5 minutes at 350g and 4°C. Washing was repeated for a total of 3 washes. Subsequently, the cells were resuspended in an appropriate volume of 1x DPBS (Gibco), passed through a 40 μm mesh (Flowmi™ Cell Strainer, Merck) and counted, using a Neubauer counting (Marienfeld). Cell counts were adjusted and hashtagged cells were pooled equally. The cell suspension was super-loaded, with 50,000 cells, in the Chromium™ Controller for partitioning single cells into nanoliter-scale Gel Bead-In-Emulsions (GEMs). Single Cell 3’ reagent kit v3.1 was used for reverse transcription, cDNA amplification and library construction of the gene expression libraries (10x Genomics) following the detailed protocol provided by 10xGenomics. Hashtag libraries were prepared according to the cell hashing protocol for 10x Single Cell 3’ Reagent Kit v3.1 provided by Biolegend, including primer sequences and reagent specifications. Biometra Trio Thermal Cycler was used for amplification and incubation steps (Analytik Jena). Libraries were quantified by Qubit™ 2.0 Fluorometer (ThermoFisher) and quality was checked using 2100 Bioanalyzer with High Sensitivity DNA kit (Agilent). Sequencing was performed in paired-end mode with a S1 and S2 flow cell (2 × 50 cycles kit) using NovaSeq 6000 sequencer (Illumina).

### BD Rhapsody single-cell RNA-seq

Whole transcriptome analyses, using the BD Rhapsody Single-Cell Analysis System (BD, Biosciences) were performed on PBMC and whole blood samples prepared as described above. Cells from each sample were labeled with sample tags (BD™ Human Single-Cell Multiplexing Kit) following the manufacturer’s protocol. Briefly, a total number of 1×10^6^ cells were resuspended in 180μl of Stain Buffer (FBS) (BD Pharmingen). The sample tags were added to the respective samples and incubated for 20 min at room temperature. After incubation, 200μl stain buffer was added to each sample and centrifuged for 5 minutes at 300g and 4°C. Samples were washed one more time. Subsequently cells were resuspended in 300μl of cold BD Sample Buffer and counted using Improved Neubauer Hemocytometer (INCYTO). Labelled samples were pooled equally in 650μl cold BD Sample Buffer. For each pooled sample two BD Rhapsody cartridges were super-loaded with approximately 60,000 cells each. Single cells were isolated using Single-Cell Capture and cDNA Synthesis with the BD Rhapsody Express Single-Cell Analysis System according to the manufacturer’s recommendations (BD Biosciences). cDNA libraries were prepared using the BD Rhapsody™ Whole Transcriptome Analysis Amplification Kit following the BD Rhapsody™ System mRNA Whole Transcriptome Analysis (WTA) and Sample Tag Library Preparation Protocol (BD Biosciences). The final libraries were quantified using a Qubit Fluorometer with the Qubit dsDNA HS Kit (ThermoFisher) and the size-distribution was measured using the Agilent high sensitivity D5000 assay on a TapeStation 4200 system (Agilent technologies). Sequencing was performed in paired-end mode (2*75 cycles) on a NovaSeq 6000 and NextSeq 500 System (Illumina) with NovaSeq 6000 S2 Reagent Kit (200 cycles) and NextSeq 500/550 High Output Kit v2.5 (150 Cycles) chemistry, respectively.

### Data pre-processing of 10x Genomics Chromium scRNA-seq data

CellRanger v3.1.0 (10x Genomics) was used to process scRNA-seq. To generate a digital gene expression (DGE) matrix for each sample, we mapped their reads to a combined reference of GRCh38 genome and SARS-CoV-2 genome, and recorded the number of UMIs for each gene in each cell.

### Data pre-processing of BD Rhapsody scRNA-seq data

After demultiplexing of bcl files using Bcl2fastq2 V2.20 from Illumina and quality control, paired-end scRNA-seq reads were filtered for valid cell barcodes using the barcode whitelist provided by BD. Cutadapt 1.16 was then used to trim NexteraPE-PE adapter sequences where needed and to filter reads for a PHRED score of 20 or above. Then, STAR 2.6.1b was used for alignment against the Gencode v33 (GRCh38.p13) reference genome. dropseq-tools 2.0.0 were used to quantify gene expression and collapse to UMI count data. For hashtag-oligo based demultiplexing of single-cell transcriptomes and subsequent assignment of cell barcodes to their sample of origin the respective multiplexing tag sequences were added to the reference genome and quantified as well.

### ScRNA-seq data analysis of 10x Chromium data of cohort 1

ScRNA-seq UMI count matrices were imported to R 3.6.2 and gene expression data analysis was performed using the R/Seurat package 3.1.4 (Butler et al., 2018; Hafemeister and Satija, 2019). Demultiplexing of cells was performed using the *HTODemux* function implemented in Seurat.

#### Data quality control

We excluded the cells based on the following criteria: more than 5% mitochondrial reads, less than 200 expressed genes or more than 6000 expressed genes. We further excluded genes that were expressed in less than five cells. In addition, mitochondrial genes have been excluded from further analysis.

#### Data integration

First, we SCTransformed (Seurat function) the data and then selected 2,000 features with largest variance among the data sets and identified integration anchors using these features. PCA (principal component analysis) was then performed on the integrated data sets, followed by a Shared Nearest Neighbour (SNN) Graph construction using PC1 to PC30 and for the k-Nearest Neighbours (KNN) Graph construction. The clustering analysis was performed using the Louvain algorithm with a resolution of 0.6. Uniform Manifold Approximation and Projection (UMAP) was utilized to visualize the cell clusters.

Data of control (healthy) samples were obtained from https://satijalab.org/seurat/vignettes.html and https://support.10xgenomics.com/single-cell-gene-expression/datasets/1.1.0/.

#### Normalization

LogNormalization (Seurat function) was applied before downstream analysis. The original gene counts for each cell were normalized by total UMI counts, multiplied by 10,000 (TP10K) and then log transformed by log10(TP10k+1).

#### Differential expression tests and cluster marker genes

Differential expression (DE) tests were performed using FindMarkers/FindAllMarkers functions in Seurat with Wilcoxon Rank Sum test. Genes with >0.25 log-fold changes, at least 25% expressed in tested groups, and Bonferroni-corrected p-values<0.05 were regarded as significantly differentially expressed genes (DEGs). Cluster marker genes were identified by applying the DE tests for upregulated genes between cells in one cluster to all other clusters in the dataset. Top ranked genes (by log-fold changes) from each cluster of interest were extracted for further illustration.

#### Cluster annotation

Clusters were annotated based on a double-checking strategy: 1) by comparing cluster marker genes with public sources, and 2) by directly visualizing the expression pattern of CyTOF marker genes.

#### GO enrichment analysis

Significantly differentially expressed genes (DEGs) between each monocyte cluster and the rest of monocyte subpopulations were identified by FindMarkers function from the Seurat package using Wilcoxon Rank Sum test statistics for genes expressed in at least 25% of all monocyte clusters. P-values were corrected for multiple testing using Bonferroni correction and genes with corrected P-values lower or equal 0.05 have been taken as significant DEGs for GO enrichment test by R package/ClusterProfiler v.3.10.1 (Yu et al., 2012).

#### Subset analysis of the neutrophils within the PBMC data set of cohort 1

The neutrophil space was investigated by subsetting the PBMC dataset to those clusters identified as neutrophils and pre-neutrophils (cluster 9 and 13). Within those subsets, we selected top 2000 variable genes and repeated a clustering using the SNN-graph based Louvain algorithm mentioned above with a resolution of 0.6. The dimensionality of the data was then reduced to 10 PCs, which served as input for the UMAP calculation. To categorize the observed neutrophil clusters into the respective cell cycle states, we applied the CellCycleScoring function of Seurat and visualized the results as pie charts.

A gene signature enrichment analysis using the ‘AUCell’ method (Aibar et al., 2017) was applied to link observed neutrophil clusters to existing studies and neutrophils of cohort 2. We set the threshold for the calculation of the area under the curve (AUC) to marker genes from collected publications and top 30 of the ranked maker genes from each of neutrophil clusters from cohort 2. The resulting AUC values were normalized the maximum possible AUC to 1 and subsequently visualized in violin plots or UMAP plots.

### ScRNA-seq data analysis of Rhapsody data of cohort 2

#### General steps for Rhapsody data downstream analysis

ScRNA-seq UMI count matrices were imported to R 3.6.2 and gene expression data analysis was performed using the R/Seurat package 3.1.2 (Butler et al., 2018). Demultiplexing of cells was performed using the *HTODemux* function implemented in Seurat. After identification of singlets, cells with more than 25% mitochondrial reads, less than 250 expressed genes or more than 5000 expressed genes were excluded from the analysis and only those genes present in more than 5 cells were considered for downstream analysis. The following normalization, scaling and dimensionality reduction steps were performed independently for each of the data subsets used for the different analyses as indicated respectively. In general, gene expression values were normalized by total UMI counts per cell, multiplied by 10,000 (TP10K) and then log transformed by log10(TP10k+1). Subsequently, the data was scaled, centered and regressed against the number of detected genes per cell to correct for heterogeneity associated with differences in sequencing depth. For dimensionality reduction, PCA was performed on the top 2000 variable genes identified using the vst method implemented in Seurat. Subsequently, UMAP was used for two-dimensional representation of the data structure. No batch effect removal or data integration analysis was performed on the BD Rhapsody data. Cell type annotation was based on the respective clustering results combined with data-driven cell type classification algorithms based on reference transcriptomes data (Aran et al., 2019) and expression of known marker genes.

#### scRNA-seq analysis of the complete BD rhapsody data set of cohort 2 including data from frozen and fresh PBMC and whole blood

ScRNA-seq count data of 162,797 cells derived from fresh and frozen PBMC samples purified by density gradient centrifugation and whole blood after erythrocyte lysis of cohort 2 (Bonn, BD Rhapsody) were combined, normalized and scaled as described above. After variable gene selection and PCA, UMAP was performed based on the first 25 principal components (PCs). No batch correction or data integration strategies were applied to the data. Separate visualization of the cells showed overlay of cells unaffected by the technical differences in sample handling. Data quality and information content was visualized as violin plots showing the number of detected genes and transcripts (UMIs) per sample handling strategy split by PBMC and granulocyte fraction.

#### scRNA-seq analysis of fresh and frozen PBMC samples

ScRNA-seq count data of 93,297 cells derived from fresh and frozen PBMC samples of cohort 2 (Bonn, BD Rhapsody) purified by density gradient centrifugation were normalized and scaled as described above. After variable gene selection and PCA, UMAP was performed and the cells were clustered using the Louvain algorithm based on the first 15 PCs and a resolution of 0.5. Cluster identities were determined by reference-based cell classification and inference of cluster-specific marker genes using the Wilcoxon rank sum test using the following cutoffs: genes have to be expressed in more than 20% of the cells of the respective cluster, exceed a logarithmic fold change cutoff to at least 0.25, and exhibited a difference of > 10% in the detection between two clusters.

#### Quantification of the percentages of cell clusters in the PBMC scRNA-seq data of both cohorts separated by disease group

To compare shifts in the monocyte and neutrophil populations in the PBMC compartment of COVID-19 patients, the percentages of the cellular subsets - as identified by clustering and cluster annotation explained above for the two independent scRNA-seq data sets (cohort 1 and cohort 2) - of the total number of PBMC in each data set were quantified per sample and visualized together in box plots. Statistical significance was inferred using *t*-test.

#### Subset analysis of the monocytes within the PBMC data set of cohort 2

The monocyte space was investigated by subsetting the PBMC dataset to those clusters identified as monocytes (cluster 1, 3, 5, 9, and 12) and repeating the variable gene selection (top 2000 variable genes), regression for the number of UMIs and scaling as described above. The dimensionality of the data was then reduced to 8 PCs, which served as input for the UMAP calculation. The SNN-graph based Louvain clustering of the monocytes was performed using a resolution of 0.48. Marker genes per cluster were calculated using the Wilcoxon rank sum test using the following cutoffs: genes have to be expressed in > 20% of the cells, exceed a logarithmic fold change cutoff to at least 0.25, and exhibited a difference of > 10% in the detection between two clusters.

#### Time kinetics analysis of identified monocyte clusters

For each patient and time point of sample collection, the proportional occupancy of the monocyte clusters was calculated, and the relative proportions were subsequently visualized as a function of time.

#### Analysis of the scRNA-seq data from fresh PBMC and whole blood samples of cohort 2

ScRNA-seq count data derived from fresh PBMC samples purified by density gradient centrifugation and whole blood after erythrocyte lysis of cohort 2 (BD Rhapsody) were normalized, scaled, and regressed for the number of UMI per cell as described above. After PCA based on the top 2000 variable genes, UMAP was performed using the first 15 PCs. Cell clusters were determined using Louvain clustering implemented in Seurat based on the first 15 principle components and a resolution of 0.5. Cluster identities were assigned as detailed above using reference-based classification and marker gene expression. Subsequently, the dataset was subsetted for clusters identified as neutrophils and pre-neutrophils, and re-scaled and regressed. After PCA on the top 2000 variable genes, UMAP was performed on the first 15 PCs. Clustering was performed as described above on the top 15 PCs using a resolution of 0.5. Clusters featuring high counts in hemoglobin genes and expression of the eosinophil specific marker gene CLC were excluded from subsequent analyses. After filtering and rescaling of the neutrophil compartment a division of the cells according to experimental batches was observed in the two-dimensional UMAP representation. Comparing experimental procedures of different experiments pointed to short-term storage of whole blood on ice for a single experimental batch as a technical influence potentially explaining the transcriptional differences. Therefore, cells from the respective experiment were excluded, reducing the number of samples to a total of 15 (30,019 cells, 10 controls, 5 COVID-19 patients) to prevent any technical bias from skewing the analysis. After exclusion, the remaining data were processed as described above and clustered using Louvain clustering based on the first 15 PCs with a resolution of 0.7 was performed.

Differentially expressed genes between clusters were defined using a Wilcoxon rank sum test for differential gene expression implemented in Seurat. Genes had to be expressed in >10% of the cells of a cluster, exceed a logarithmic threshold >0.1 and to have >10% difference in the minimum detection between two clusters.

#### Quantification of the percentages of cell subsets in the fresh whole blood scRNA-seq data of cohort 2

After cell type classification of the combined scRNA-seq data set of fresh PBMC and whole blood samples of cohort 2 described above, 69.500 cells derived from whole blood samples after erythrocyte lysis were subsetted. Percentages of cell subsets in those whole blood samples of the total number of cells were quantified per sample and visualized r in box plots separated by disease stage and group.

#### Confusion matrix

For each cluster of neutrophils, the relative proportion across disease severity and time point was visualized as a fraction of samples from the respective condition contributing to the cluster.

#### GO enrichment

Gene set enrichment was performed on gene sets from the Kyoto Encyclopedia of Genes and Genomes (KEGG) database (Kanehisa, 2019), Hallmark gene sets (Liberzon et al., 2015) and Gene Ontology (GO) (Ashburner et al., 2000; Carbon et al., 2019) using the R package/ClusterProfiler v.3.10.1 (Yu et al., 2012).

#### Cell cycle state analysis of scRNA-Seq data

To categorize the cells within the neutrophil clusters into the respective cell cycle states, we applied the *CellCycleScoring* function of Seurat and visualized the results as pie charts.

#### Trajectory analysis

Trajectory analysis was performed using the *destiny* algorithm (Angerer et al., 2016). In brief, the neutrophil space was subsetted to only severe patients (early and mild) and only the most prominent clusters of the latter (clusters 9,8,2,0,6). The normalized data were scaled and regressed for UMIs and a diffusion map was calculated based on the top 2.000 variable genes with a sum of at least 10 counts over all cells. Based on the diffusion map, a diffusion pseudo time was calculated (without fixing a starting point) to infer a transition probability between the different cell states of the neutrophils. Subsequently, the density of the clusters along the pseudotime and marker gene expression for each cluster were visualized.

Enrichment of gene sets was performed using the ‘AUCell’ method (Aibar et al., 2017) implemented in the package (version 1.4.1) in R. We set the threshold for the calculation of the AUC to the top 3% of the ranked genes and normalized the maximum possible AUC to 1. The resulting AUC values were subsequently visualized in violin plots or UMAP plots.

#### Transcription factor prediction analysis

The Cytoscape (version v3.7.1, doi: 10.1101/gr.1239303) plug-in iRegulon (Janky et al., 2014) (version 1.3) was used to predict the transcription factors potentially regulating cluster-specifically expressed gene sets in the neutrophil subset analysis in cohort 2. The genomic regions for TF-motif search were limited to 10 kbp around the respective transcriptional start sites and filtered for predicted TFs with a normalized enrichment score > 4.0. Next, we filtered for TFs, which exceeded a cumulative normalized expression cutoff of 50 in the respective neutrophil cluster. Subsequently, we selected transcription factors of known relevance in the context of neutrophil biology and constructed a network linking target genes among the cluster-specifically expressed marker genes and their predicted and expressed regulators for visualization in Cytoscape.

### Mass cytometry data analysis

Cytobank.org was used for de-barcoding of individual samples and manually gating of cell events to remove doublets, normalization beads and dead cells (Kotecha et al., 2010). For semi-automated gating of populations of interest, high-resolution SPADE clustering was conducted on all indicated markers (supplementary table 2) with 400 target nodes. Individual SPADE nodes were then aggregated and annotated to cell subsets (bubbles) according to the expression of lineage-specific differentiation markers.

To generate tSNE maps viSNE analysis was performed using the indicated markers (supplementary table 2). Embedding parameters were set to at least 1000 iterations per 100.000 analyzed cells, perplexity of 30 and theta of 0.5. For statistical analysis of cell population abundances, we fitted a generalized linear mixed-effects model (GLMM) for each population using the lme4 package as previously described by Nowicka et. al (Robinson et al., 2017).

## Data Availability

Data are deposited at the European Genome-phenome Archive (EGA) under access number EGAS00001004450, which is hosted by the EBI and the CRG.

## Data Analysis of Flow Cytometry Data

Flow cytometry data analysis was performed with FlowJo V10.6.1. Relative cell percentage or mean fluorescence intensity (MFI) was used for visualization and statistical analysis. Cell type was defined as granulocytes (CD45^+^, CD66b^+^), non-classical monocytes (CD45^+^, CD66b^-^, CD19^-^, CD3^-^, CD56^-^, CD14^dim^, CD16^+^).

## Data visualization

In general, the R packages Seurat and the ggplot2 package (version 3.1.0, Wickham, 2016) were used to generate figures. For visualization of mass cytometry data, cluster minimum-spanning trees were rendered using Cytobank, the ComplexHeatmap package was used to display subset phenotypes and GraphPad Prism to generate boxplots of quantitative data.

## Lead Contact

Further information and requests for resources and reagents should be directed to the Lead Contact, **Joachim L. Schultze**

## KEY RESOURCES TABLE

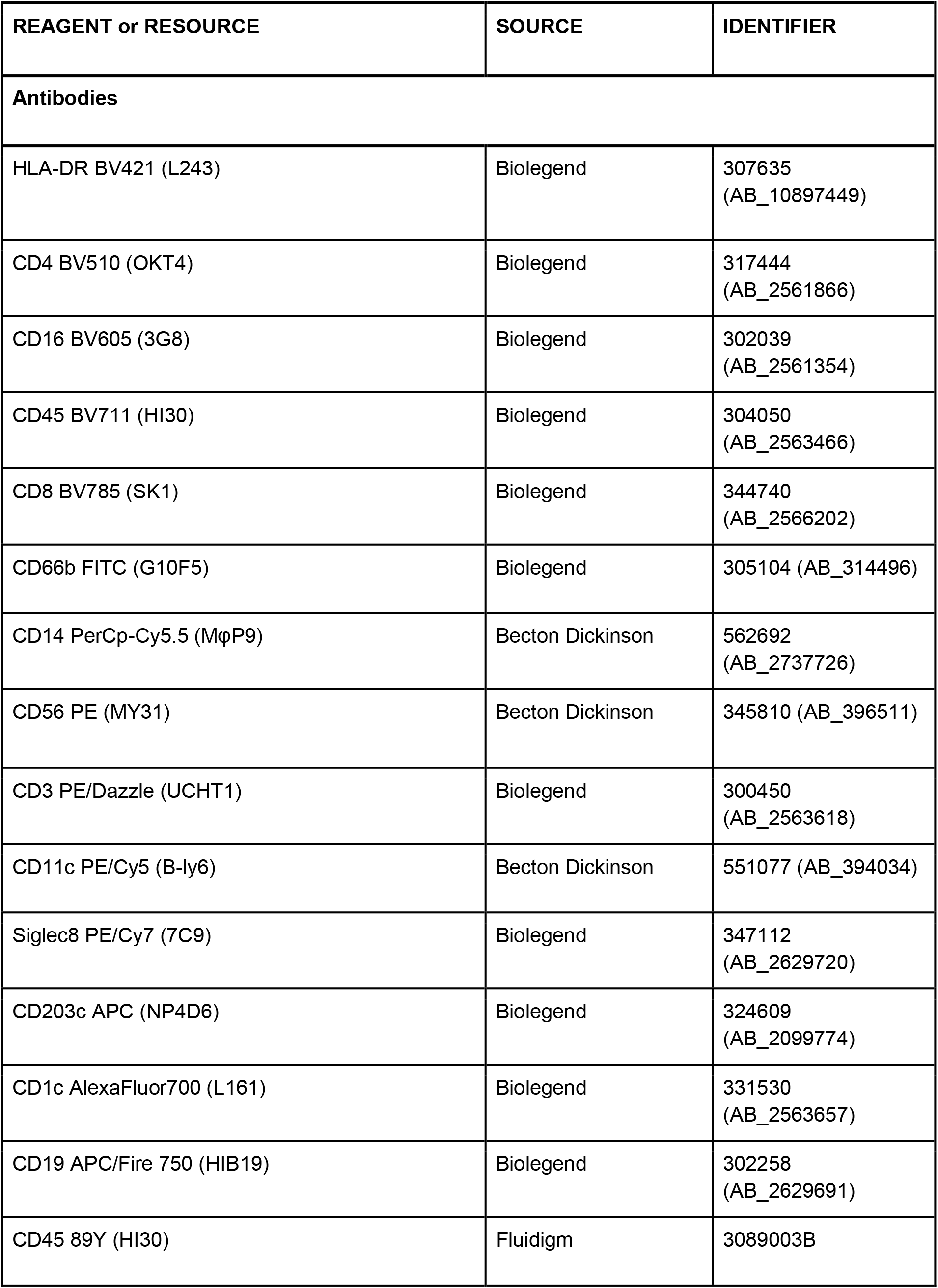
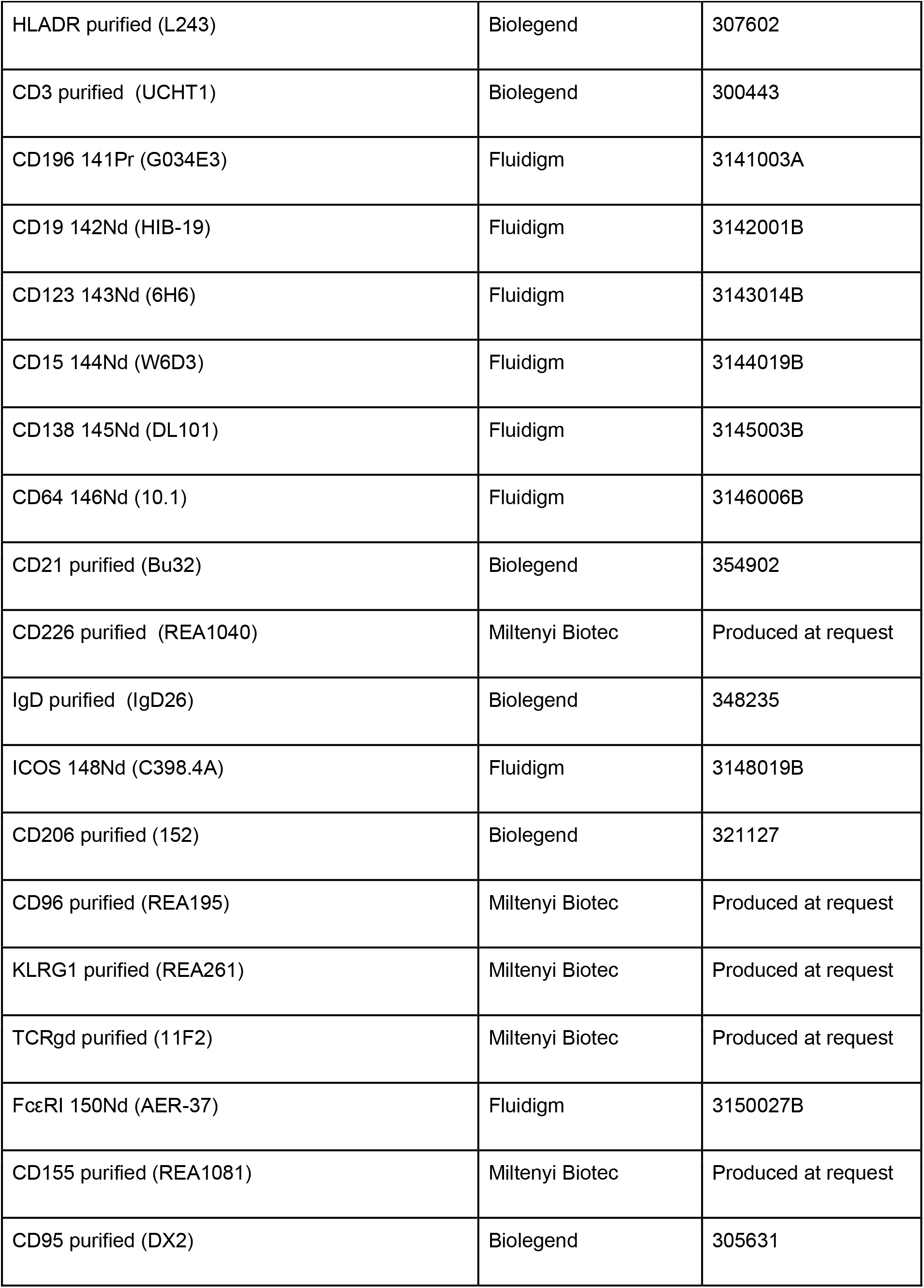
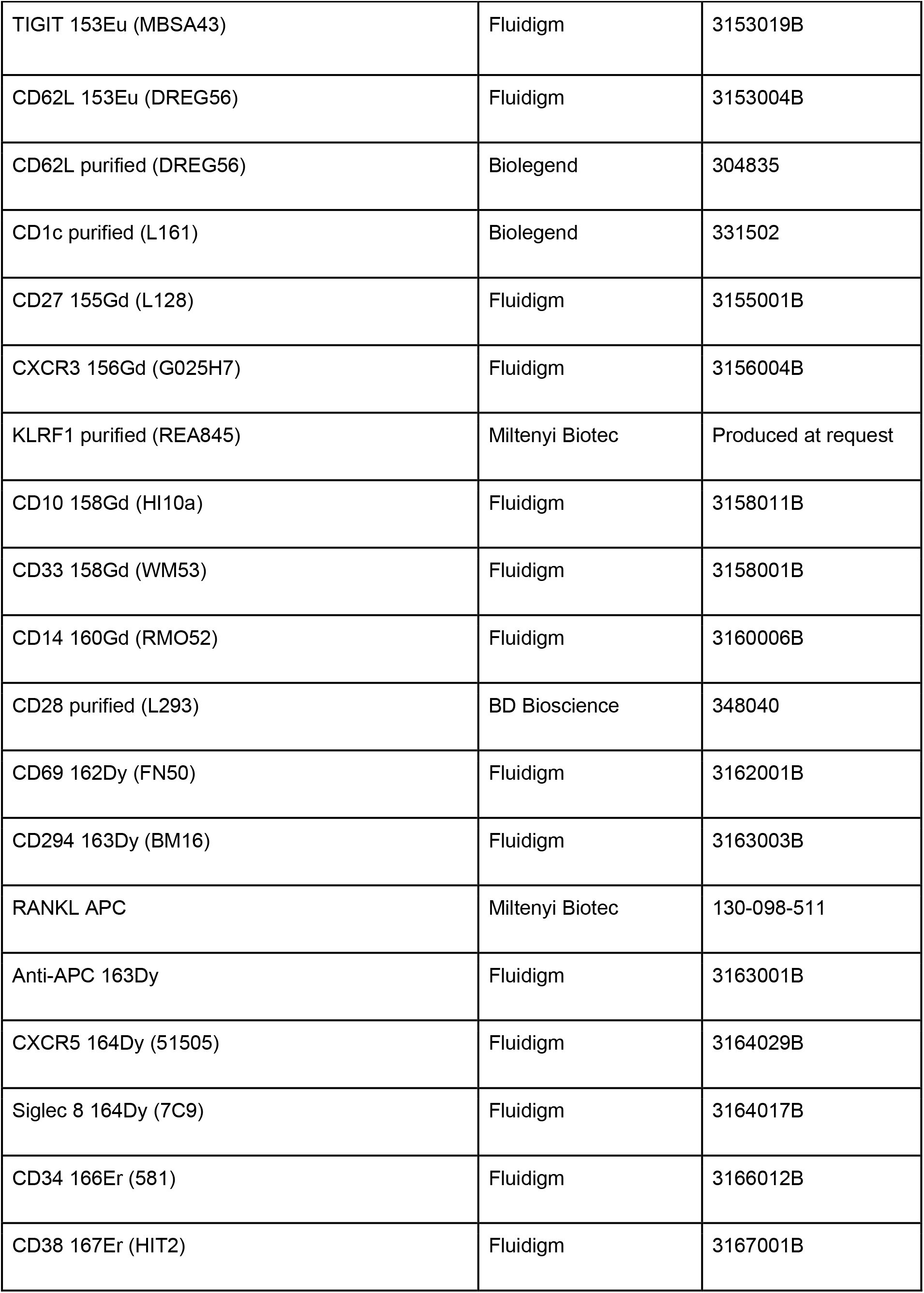
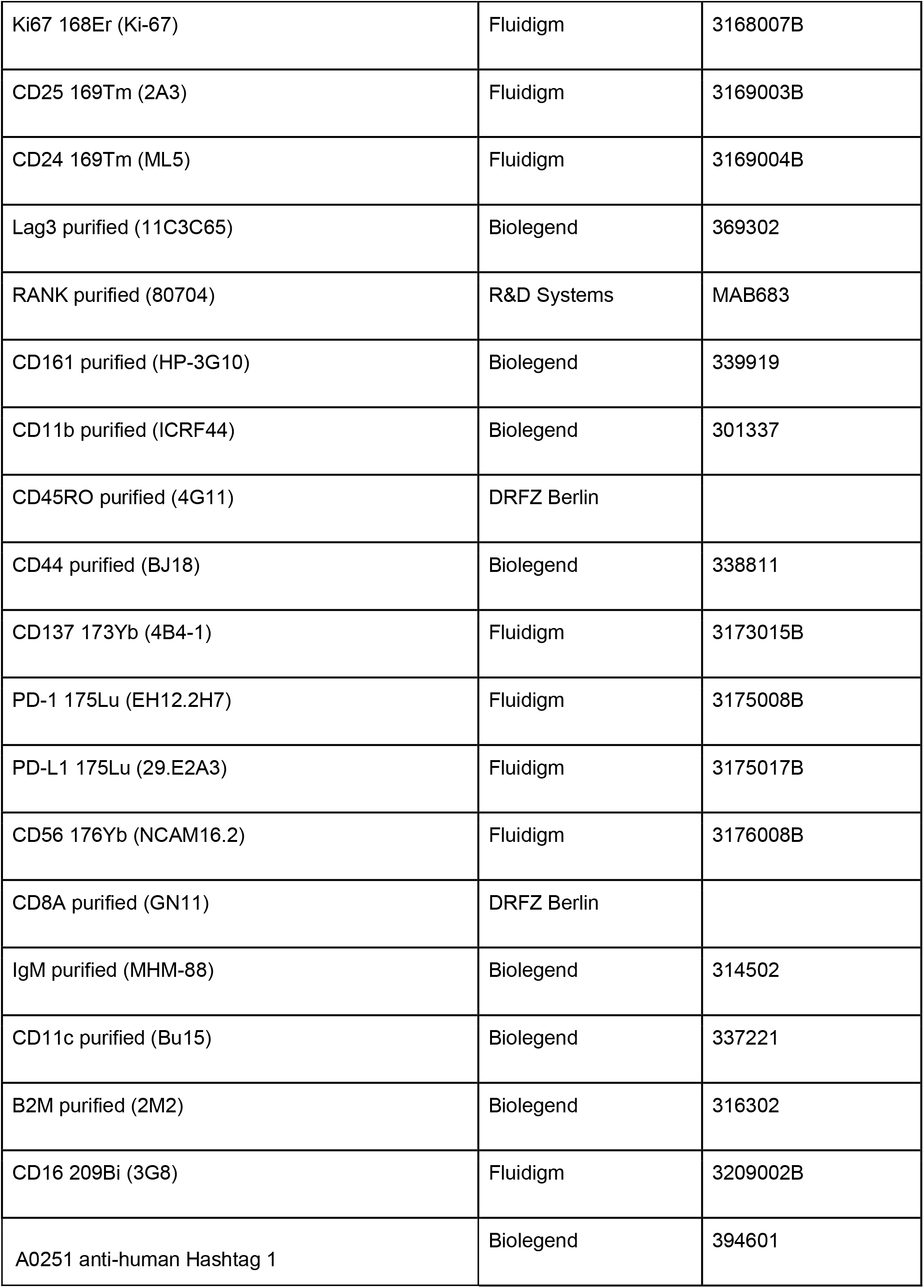
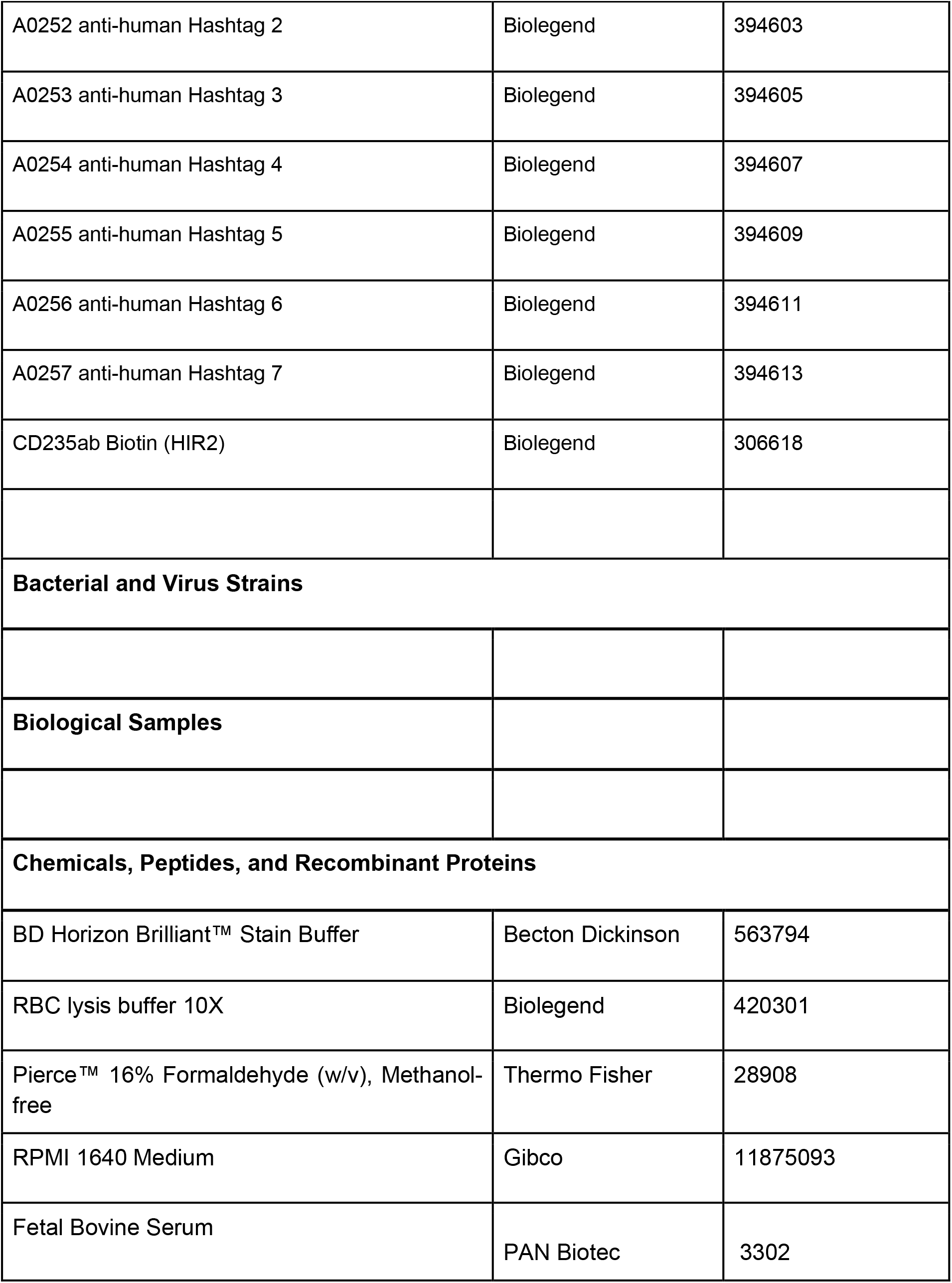
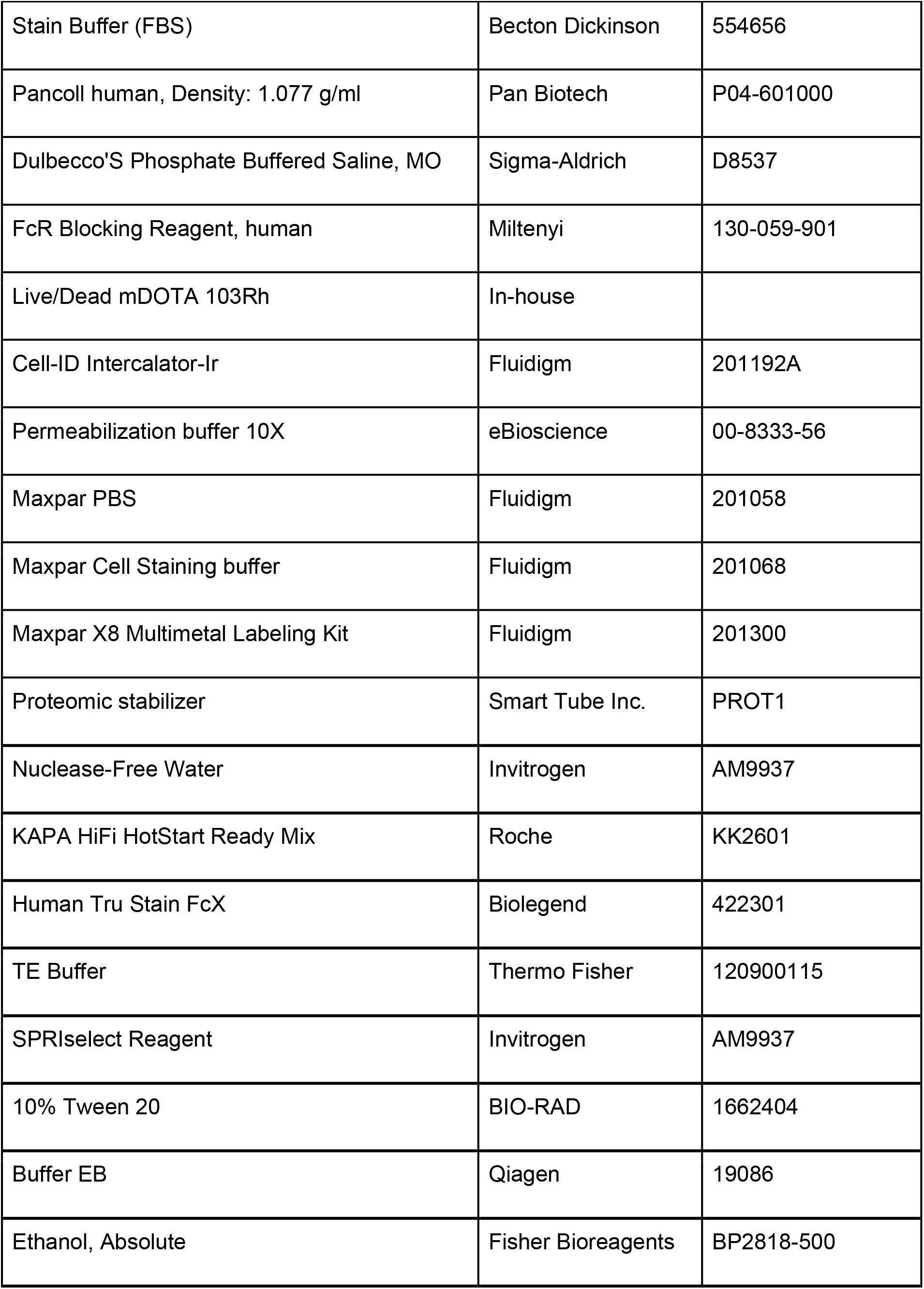
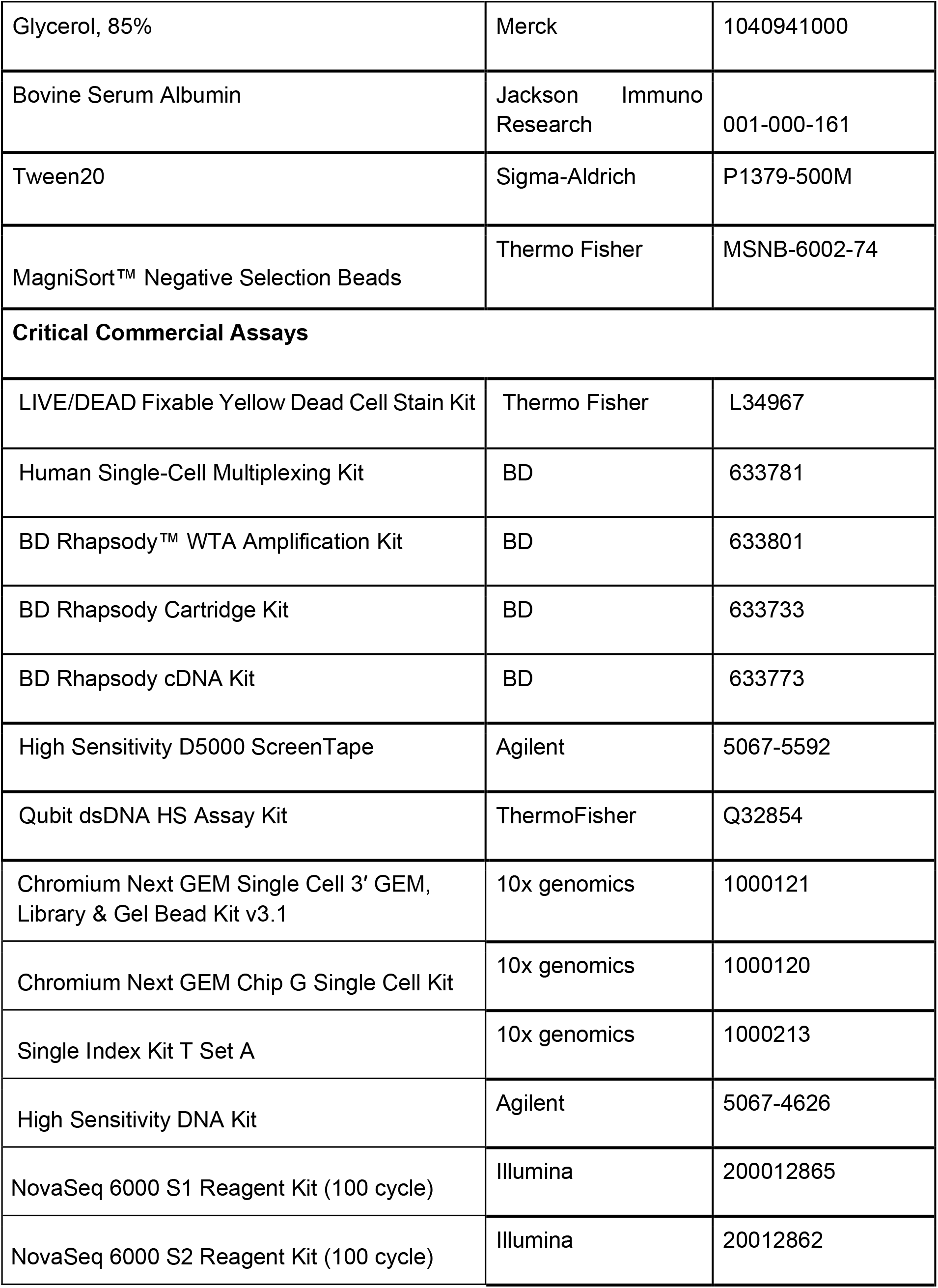
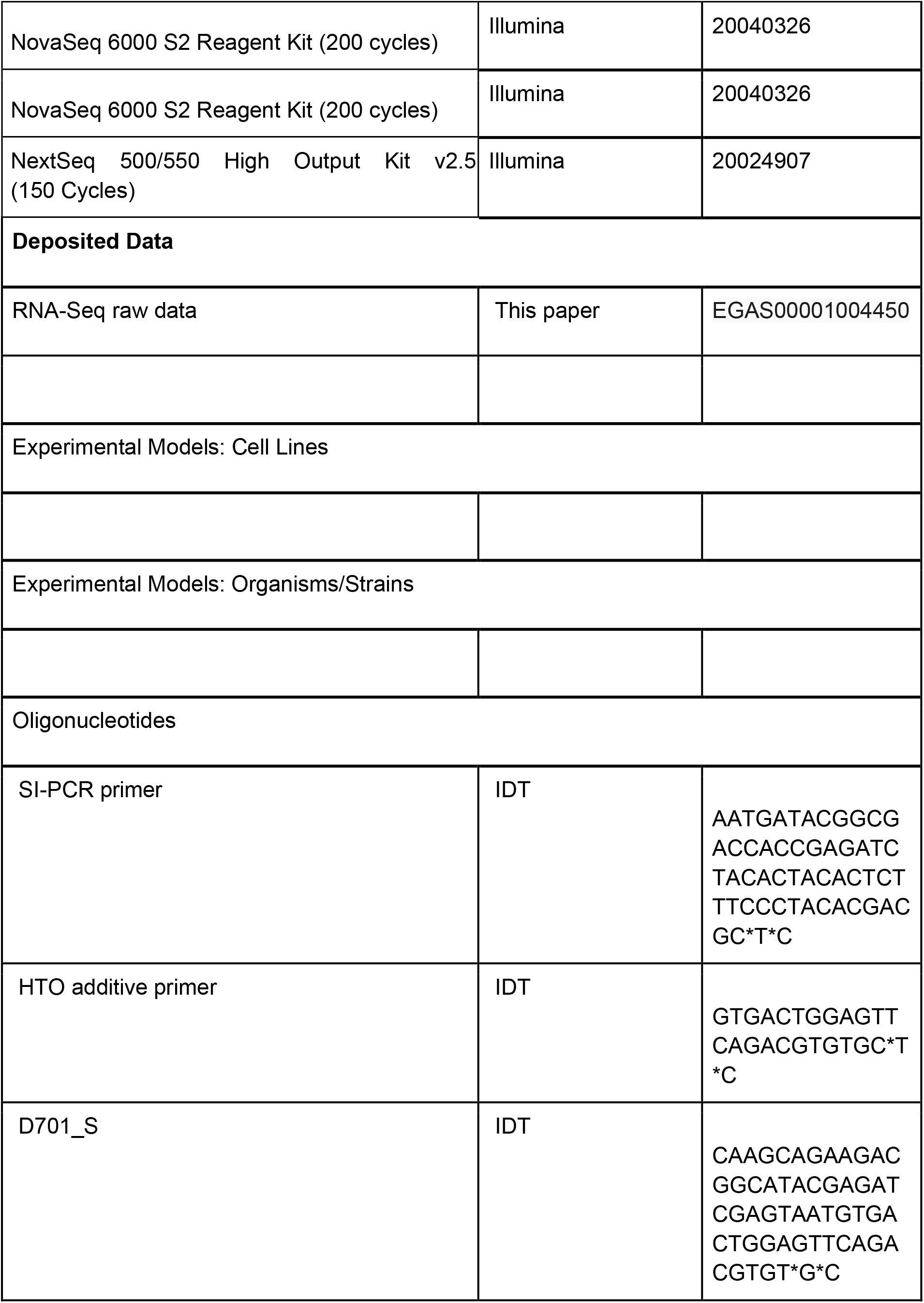
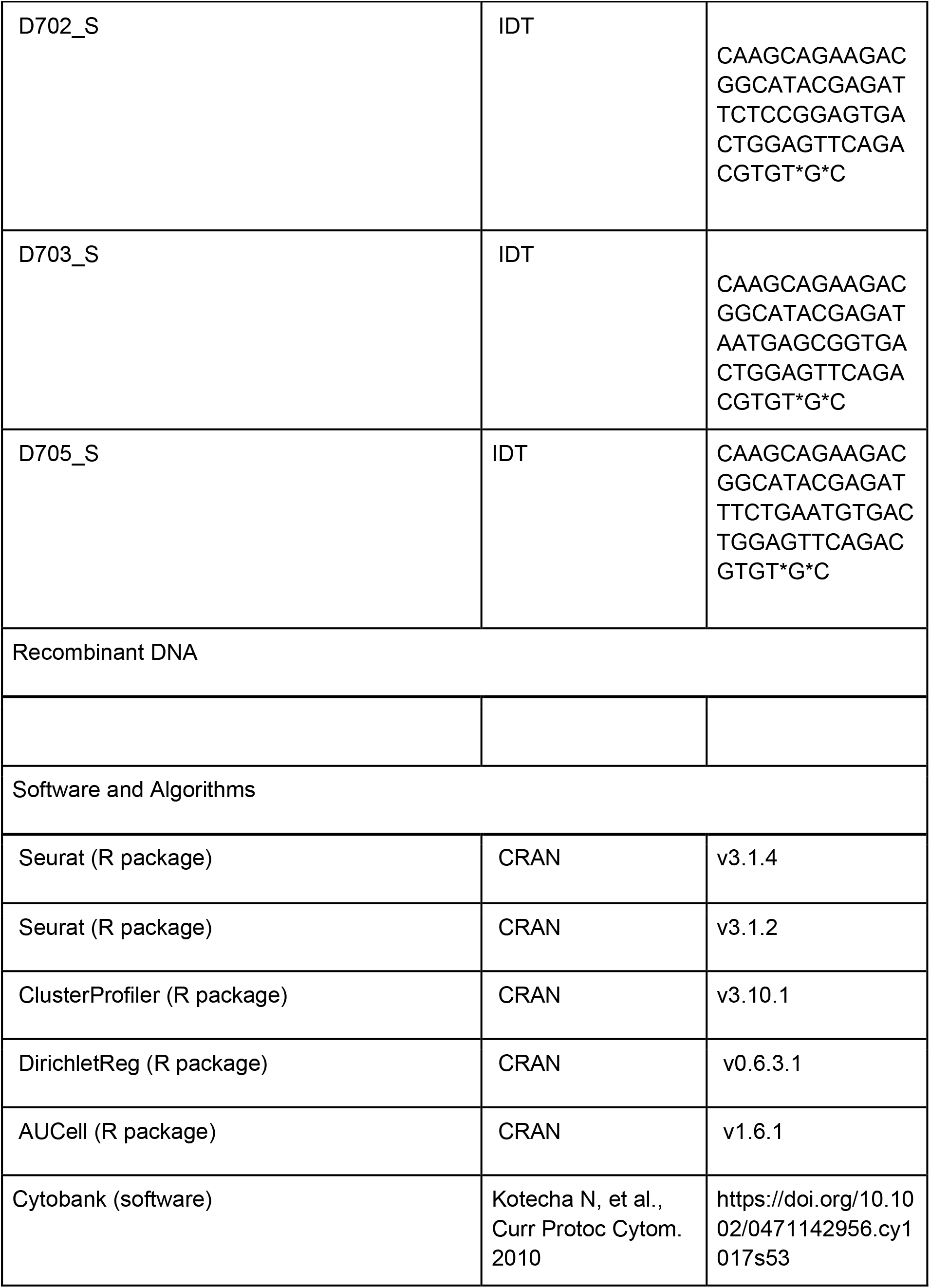
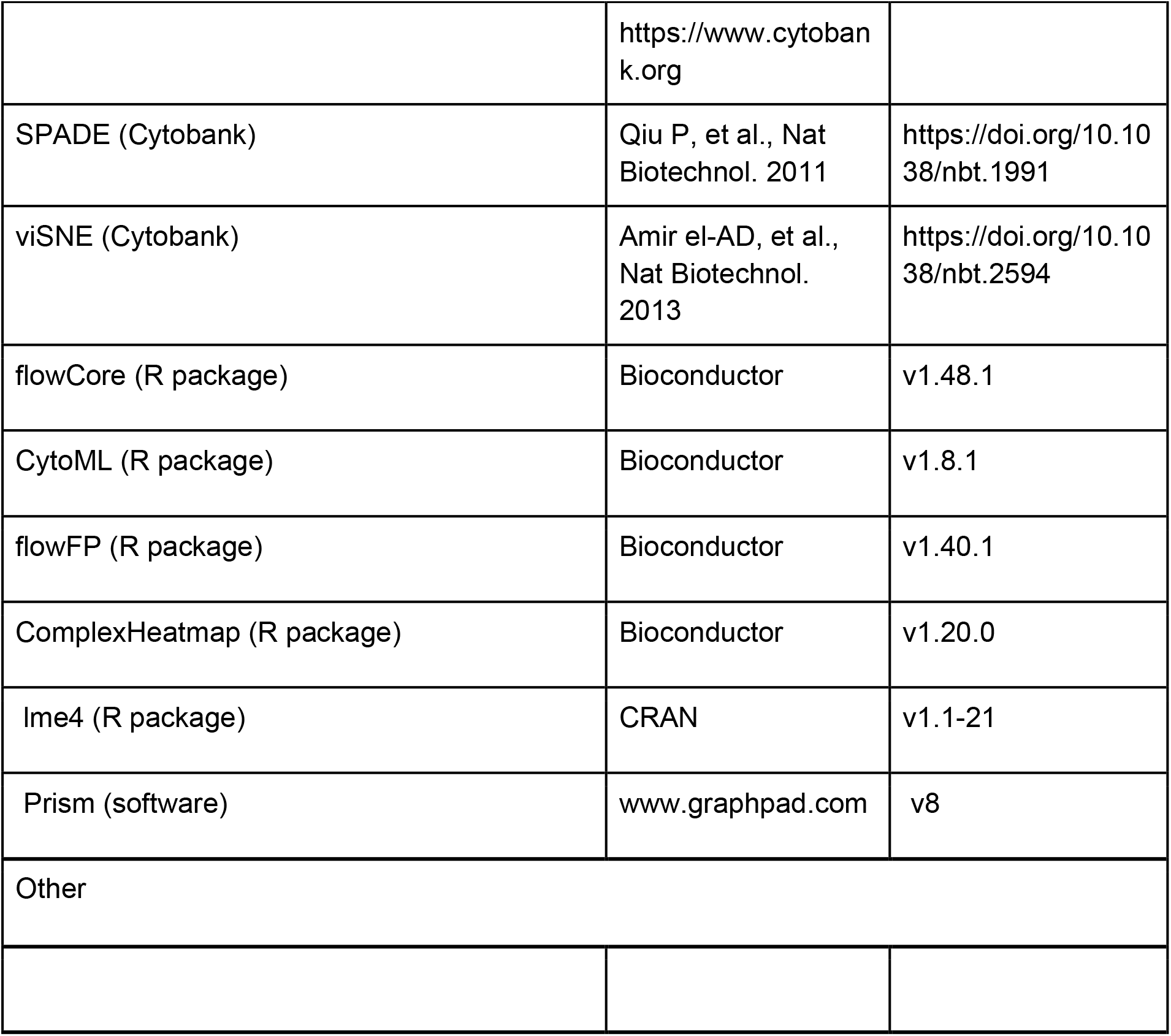

## Acknowledgements

We thank Michael Kraut and Heidi Theis for technical assistance.

## Deutsche COVID-19 Omics Initiative (DeCOI)

Robert Bals, Alexander Bartholomäus, Anke Becker, Ezio Bonifacio, Peer Bork, Thomas Clavel, Maria Colome-Tatche, Andreas Diefenbach, Alexander Dilthey, Nicole Fischer, Konrad Förstner, Julien Gagneur, Alexander Goesmann, Torsten Hain, Michael Hummel, Stefan Janssen, René Kallies, Birte Kehr, Andreas Keller, Sarah Kim-Hellmuth, Christoph Klein, Oliver Kohlbacher, Jan Korbel, Ingo Kurth, Markus Landthaler, Yang Li, Kerstin Ludwig, Oliwia Makarewicz, Manja Marz, Alice McHardy, Christian Mertes, Markus Nöthen, Peter Nürnberg, Uwe Ohler, Stephan Ossowski, Jörg Overmann, Klaus Pfeffer, Alfred Pühler, Nikolaus Rajewsky, Markus Ralser, Olaf Rieß, Stephan Ripke, Ulisses Nunes da Rocha, Philip Rosenstiel, Antoine-Emmanuel Saliba, Leif Erik Sander, Birgit Sawitzki, Philipp Schiffer, Joachim L. Schultze, Alexander Sczyrba, Oliver Stegle, Jens Stoye, Fabian Theis, Janne Vehreschild, Jörg Vogel, Max von Kleist, Andreas Walker, Jörn Walter, Dagmar Wieczorek, John Ziebuhr

## Funding

This work was supported by the German Research Foundation (DFG, SFB-TR84 #114933180 to L.E.S., S.H., A.H., N.S., C.D.; INST 37/1049-1, INST 216/981-1, INST 257/605-1, INST 269/768-1 and INST 217/988-1, INST 217/577-1 to J.L.S; SFB TR57, SPP1937 to J.N.; GRK2157 to A.-E.S., ME 3644/5-1 to H.E.M.); the Berlin University Alliance (BUA, PreEP-Corona grant to L.E.S., V.C. and C.D.), the Berlin Institute of Health (BIH, grant to L.E.S., A.H., C.D.); the HGF grant sparse2big, the EU projects SYSCID (grant number 733100), and ERA CVD (grant number 00160389) to J.L.S.; the DZIF (TTU 04.816, 04.817 to J.N.); the Hector-Foundation (M89 to J.N.); the EU projects ONE STUDY (grant number 260687 to B.S.), BIO-DrIM (305147 to B.S.) and INsTRuCT (grant number 860003 to B.S.), the German Federal Ministry of Education and Research (BMBF, project RAPID to C.D., S.H., A.H.), and a Charité ^3^R project to B.S. and S.H.; Radboud University Medical Centre Hypatia Grant (2018 to Y.L.).

## Declaration of interests

The authors do not declare any conflict of interest

## Author contributions

Conceptualization: J. S-S., N.R., K.B., B.K., L.B., F.K., J.L.S, A.C.A., Y.L., J.N., B.S., A.-E.S., L.E.S. Methodology: J. S-S., D.P., T.K., S.B., L.B., E.D.D., M.G., D.W., M.B., T.S.K., A.S., O.D., H.M., A.R.S, C.C., D.K., E.V., C.J.X., A.D., C.T., S.H., C.L.G., T.U., M.B., R.G., C.D., C.V.K., K.H. Software/data analysis:J. S-S., N.R., K.B., S.S., B.Z., T.K., L.B., A.S., T.U., M.B. Investigation: J. S-S., K.B., T.P., A.H., M.H., J.L.S, A.C.A., M.W., Y.L., J.N., B.S., A.-E.S., L.E.S. Biospecimen/ enzyme resources: B.K., S.B., M.P., S.H., H.M.R., F.M., A.U., L.B.J., L.J., C.R.G., J.R., K.M.K, M.T., G.R., F.K., J.N., M.W. Writing - Original Draft: J. S-S., N.R., K.B., L.B., E.D.D., C.M., J.L.S, A.C.A., Y.L., J.N., B.S., A.-E.S., L.E.S. Writing - Review & Editing: J. S-S., N.R., M.W., K.B., L.B., E.D.D., T.P., M.B., T.S.K., S.H., A.H., M.S., H.D.V., C.D., N.S., C.V.K., F.K., J.L.S, A.C.A., Y.L., J.N., B.S., A.-E.S., L.E.S.

---

**Supplemental Figure 1.**
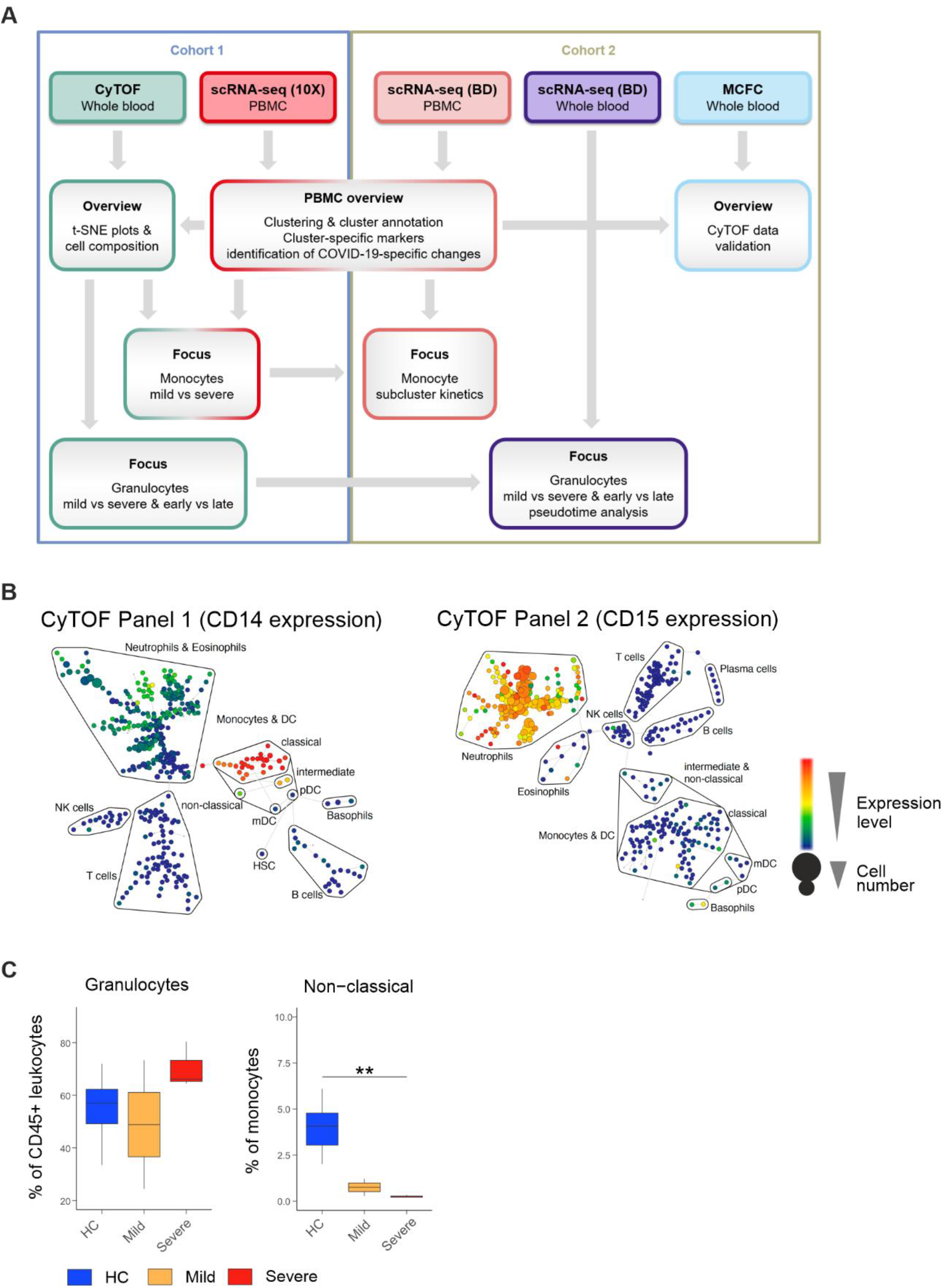
Overview of sample analysis pipeline, major leukocyte lineages definition and quantification by CyTOF and MCFC. **A**, Overview summarising sample analysis pipeline for single cell transcriptomics and proteomics of COVID-19 samples. **B**, High resolution SPADE analysis with 400 target nodes and individual nodes aggregated to the indicated major immune cell lineages according to the expression of lineage specific cell marker such as CD14 for monocytes and CD15 for neutrophils of whole blood samples collected from COVID-19 patients and healthy controls and stained with CyTOF panel 1 and 2, respectively. **C**, Box plots summarising differences in major immune cell lineage subtype composition of whole blood samples from the second cohort of COVID-19 patients showing either mild (n=2) or severe/critical disease (n=3) course, age-matched healthy controls (n=7) measured by conventional fluorochrome-based cytometry. **p<0.01.

**Supplemental Figure 2.**
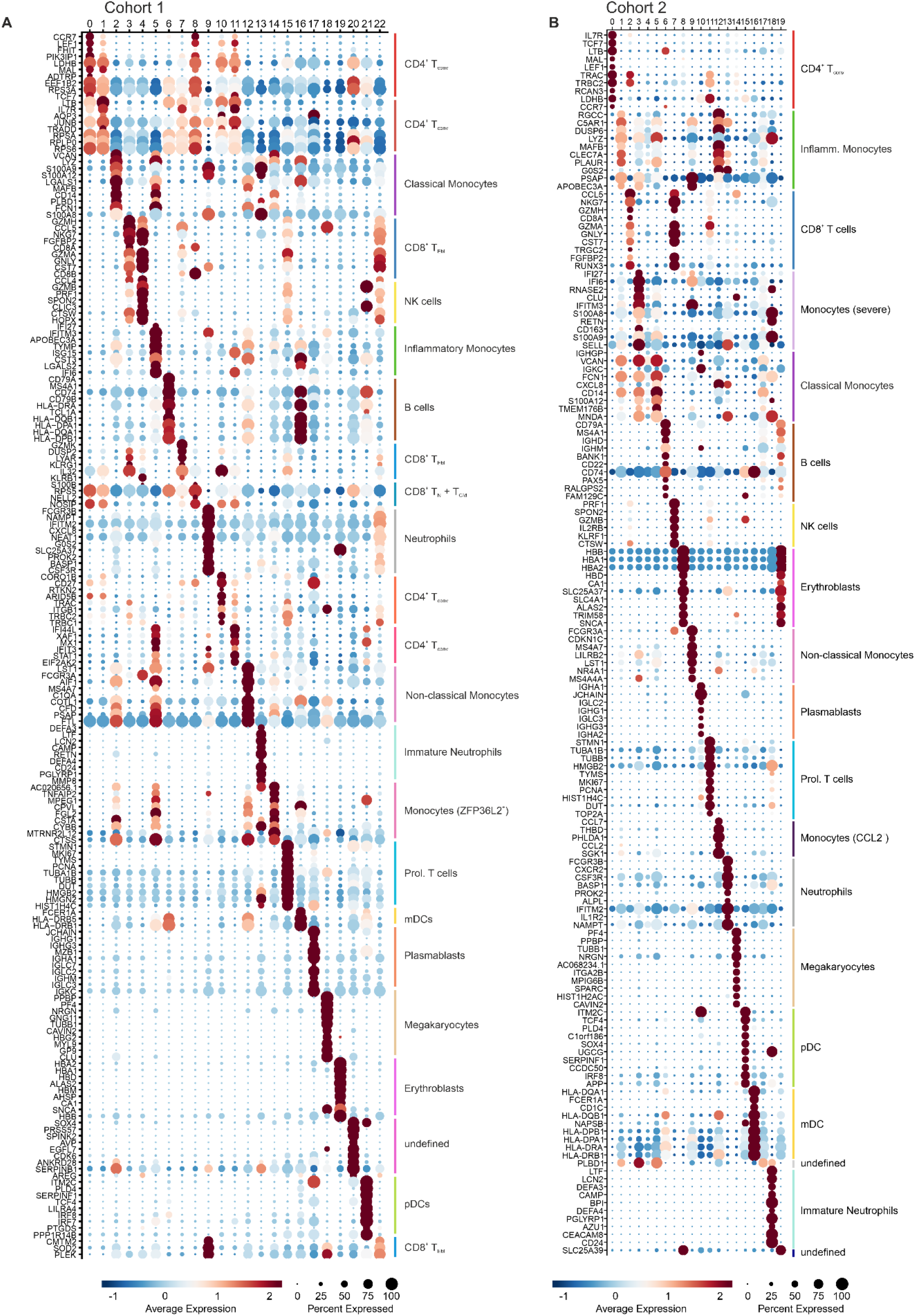
Cluster-specific marker gene expression shows inflammatory activation signatures of monocyte subsets and the appearance of neutrophil subsets in the PBMC fraction. **A**, Dot plot representation of the top 10 marker genes sorted by average log fold change determined for the clusters depicted in the UMAP in Figure 2A. **B**, Dot plot representation of the top 10 marker genes sorted by average log fold change determined for the clusters depicted in the UMAP in Figure 2C.

**Supplemental Figure 3.**
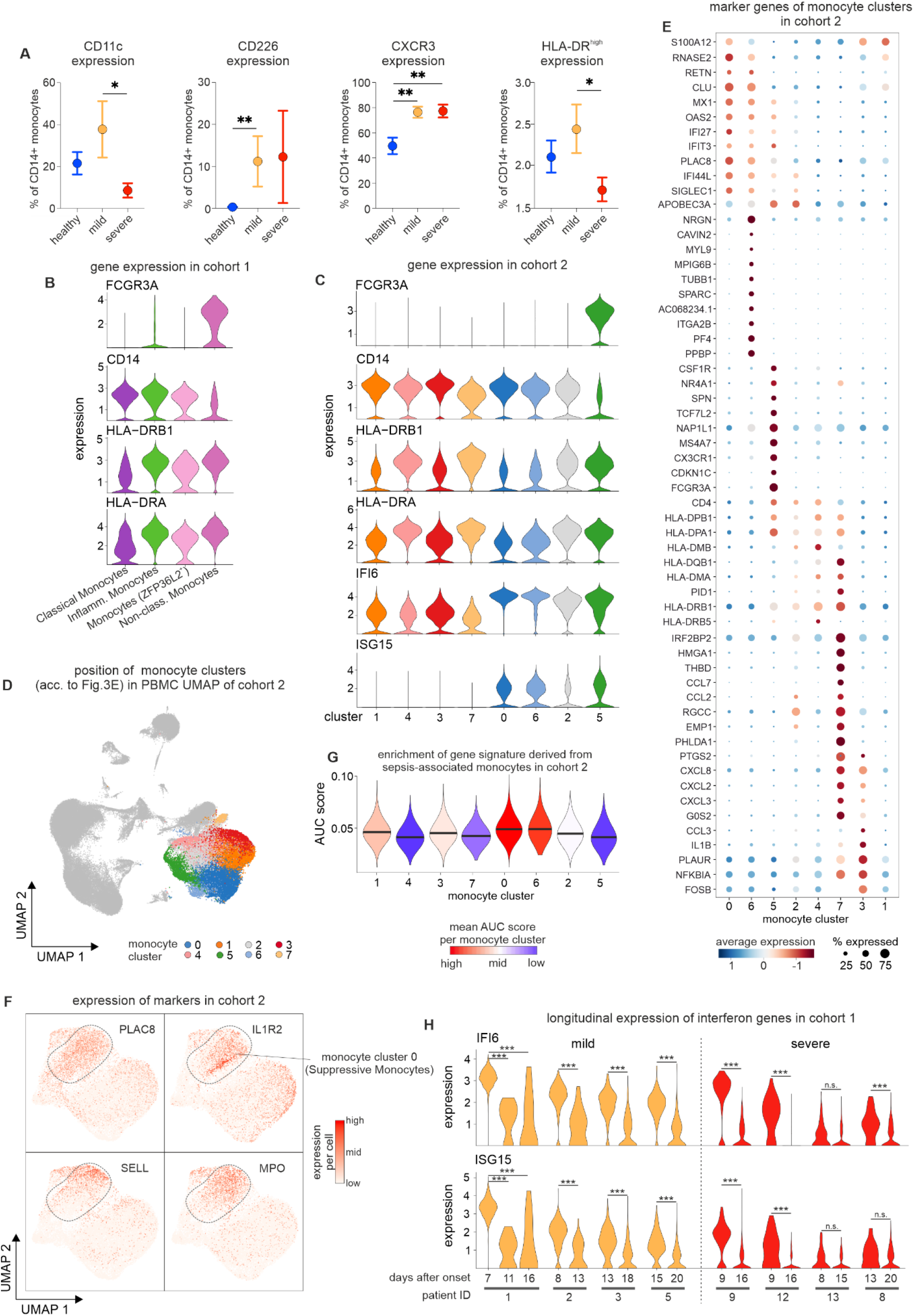
Phenotypical and transcriptional differences of monocytes from mild and severe COVID-19. **A**, Box plots summarizing differences in CD226, HLA-DRhigh, CD11c and CXCR3 expression within classical monocytes measured by mass cytometry in whole blood samples from cohort 1 distinguishing between healthy controls (n=7), COVID-19 patients with mild (n=5) or severe/critical disease (n=6) course. *p<0.05, **p>0.01 **B**, Violin plots representing expression of selected genes in monocyte clusters of cohort 1. Coloring according to cluster color in Figure 2 **C**, Violin plots representing expression of selected genes in monocyte clusters of cohort 2. **D**, Back mapping of identified monocyte clusters of cohort 2 onto the PBMC UMAP in Figure 2. **E**, Dot plot representation of marker genes found in monocyte clusters of cohort 2. **F**, Visualization of the expression of selected genes in the UMAP of the monocyte space in cohort 2. **G**, AUCell-based enrichment of gene signature derived from sepsis-associated monocytes (Reyes et al., 2020) in monocytes of cohort 2 and plotting of the ‘Area Under the Curve’ (AUC) scores as violin plots. The horizontal lines in the violin plots represent the median of the respective AUC scores per cluster. **H**, Time-dependent change of IFI6 and ISG15 expression in monocytes of cohort 1. Significant changes determined by the Wilcoxon rank sum test are indicated (= pvalue < 0.001; n.s. = not significant).

**Supplemental Figure 4.**
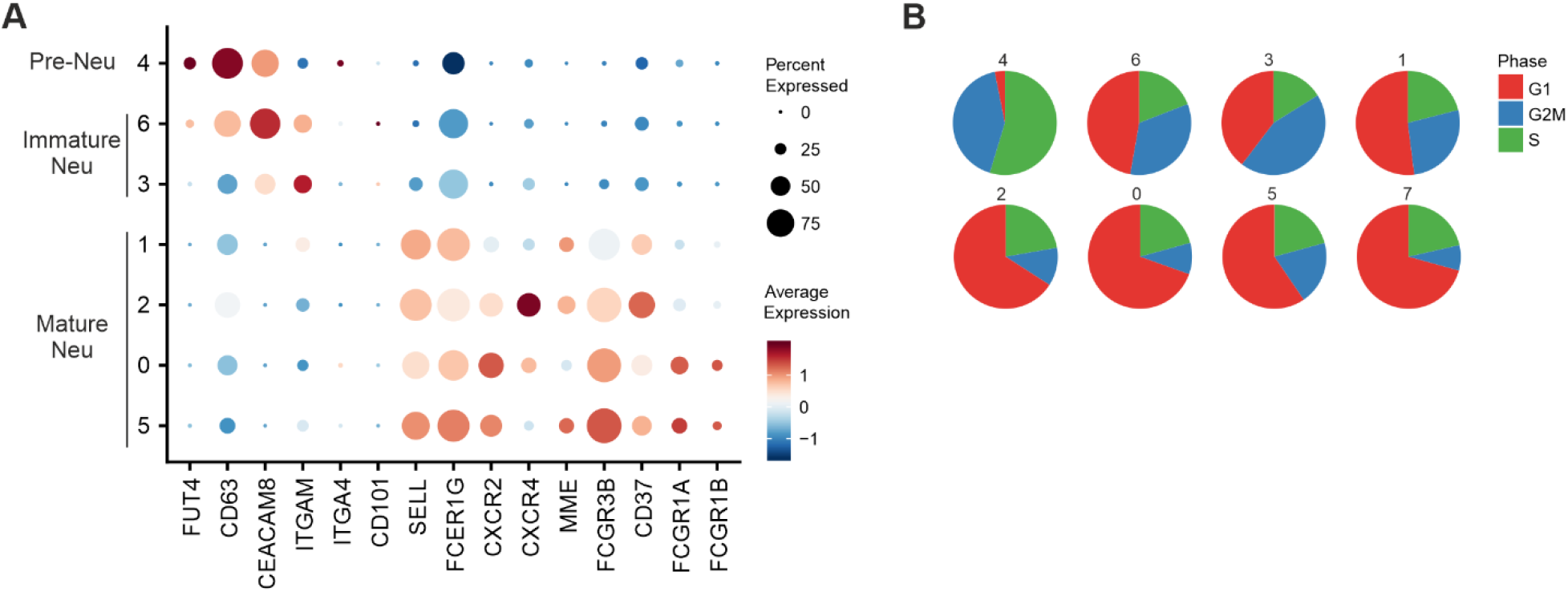
Additional analysis of dysfunctional neutrophils in PBMC fraction. **A**, Dot plot representation of marker genes associated to pre-neutrophils (pre-neutr), immature and mature neutrophils. **B**, Pie charts giving the proportion of cells in each cell cycle stage. The numbers refer to clusters as identified in the UMAP in Figure 4A.

**Supplemental Figure 6.**
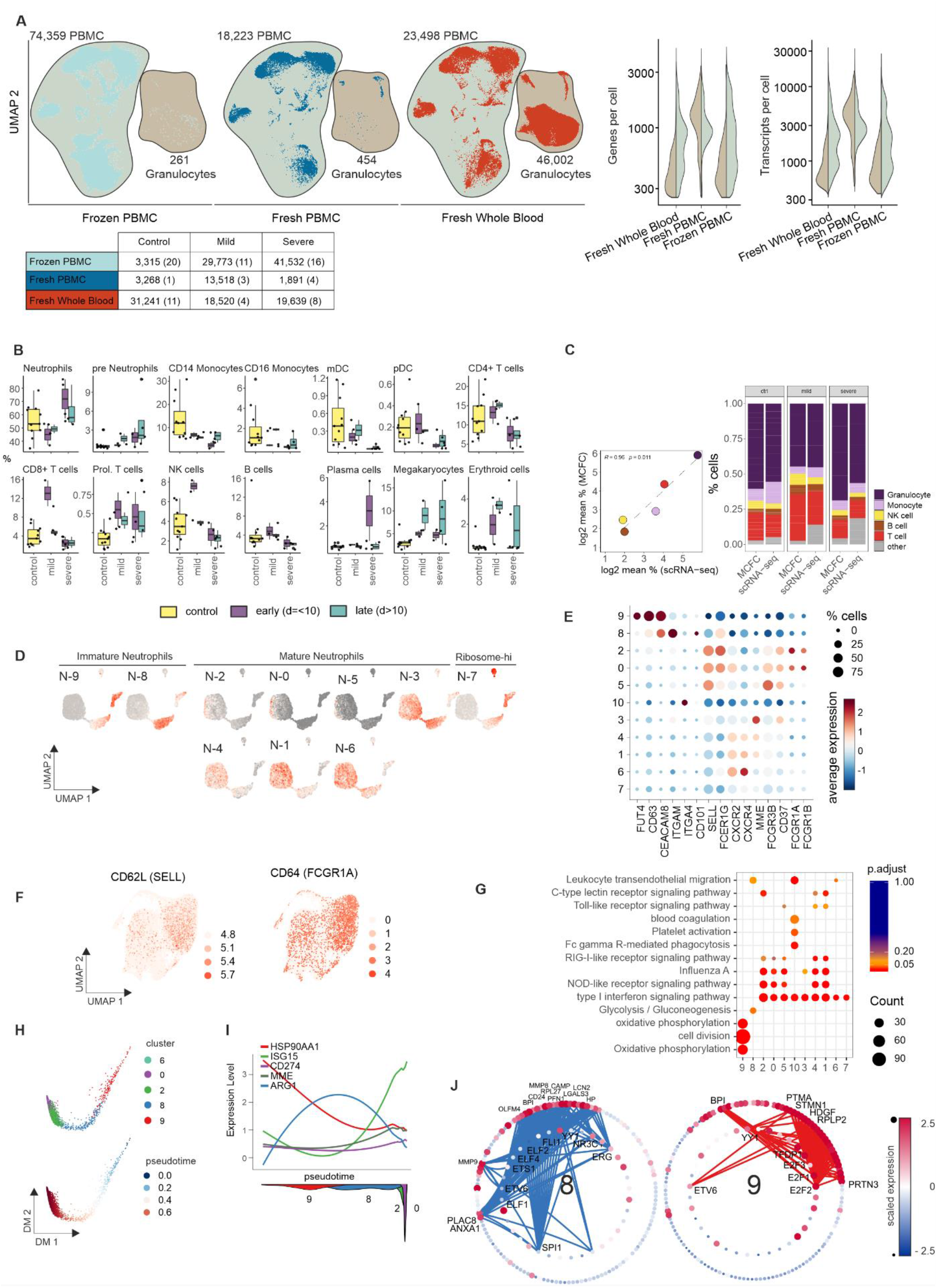
Overview of scRNA-seq dataset from cohort 2 and additional characterization of suppressive neutrophils. **A**, UMAP (right) of the complete scRNA-seq dataset from cohort 2 (frozen PBMC, fresh PBMC, fresh whole blood) and violin plot of number of genes and transcripts expressed in the PBMC VS. granulocyte fraction across the different data sets of cohort 2 (right). Violin plots are split into granulocytes (left) and PBMC (right). The table below indicates the number of cells per experimental condition with the number of samples in brackets. **B**, Box plot of cell type frequencies identified by scRNA-seq in fresh whole blood samples after erythrocyte lysis comparing 23 samples from 11 control individuals, 4 samples from mild and 8 samples from severe COVID-19 patients at early and late timepoints. **C**, Comparison between cell frequencies identified by scRNA-seq and MCFC, pearson’s correlation between the mean of each cell population (left) and stacked bar chart sorted by disease severity. **D**, Enrichment of signature genes from the neutrophil clusters from cohort 2 on the UMAP visualization of cohort 1. **E**, Dot plot representation of marker genes taken from literature classifying different neutrophil subsets. **F**, UMAP representation of neutrophils showing the scaled expression of CD62L *(SELL)* and CD64 *(FCGR1A)* with a clear enrichment in the COVID-19 specific clusters 2 and 0. **G**, Dot plot visualization of selected significantly enriched Gene Ontology terms and KEGG pathways for each cluster from the neutrophil space. **H**, Diffusion map dimensionality reduction of the main neutrophil clusters 9, 8, 2, 0 and 6 from the severe COVID-19 patients (top) and diffusion pseudotime visualized on the diffusion map indicating the transition probability of the different clusters in the following order: 9 - 8 - 2 - 0 - 6 (bottom). **I**, Genes specific for each cluster visualized along the diffusion pseudotime (top) with the density of each cluster along the pseudotime (bottom) highlighting the proposed order of differentiation of the different neutrophil subsets. **H**, Transcription factor binding prediction results for clusters 8 and 9 shown as networks of transcription factors and their targets among the specifically expressed genes for the given cluster. Edges represent predicted transcriptional regulation. Transcription factors in the inner circle and their predicted target genes in the outer circle are represented as nodes sized and colored according to the scaled expression level across all clusters. Top 10 highest connected transcription factors and those exclusively predicted for the cluster as well as the top 5 highest connected target genes together with a literature-based selection of targets are labeled.

## Supplemental Tables

**Supplemental Table 1**. Cohort outline used to perform scRNA-seq, mass cytometry and MCFC (multi-colour flow cytometry).

**Supplemental Table 2**. Detailed information on antibody panels used for mass cytometry analysis.

**Supplemental Table 3**. List of antibodies used for MCFC.

## Notes

### Competing Interest Statement

The authors have declared no competing interest.

### Author Declarations

The study was approved by the Institutional Review board of Charité (EA2/066/20)and the Institutional Review board of the University Hospital Bonn (073/19 and 134/20).

